# Genome-wide analyses of ADHD identify 27 risk loci, refine the genetic architecture and implicate several cognitive domains

**DOI:** 10.1101/2022.02.14.22270780

**Authors:** Ditte Demontis, G. Bragi Walters, Georgios Athanasiadis, Raymond Walters, Karen Therrien, Leila Farajzadeh, Georgios Voloudakis, Jaroslav Bendl, Biao Zeng, Wen Zhang, Jakob Grove, Thomas D. Als, Jinjie Duan, F. Kyle Satterstrom, Jonas Bybjerg-Grauholm, Marie Bækved-Hansen, Olafur O. Gudmundsson, Sigurdur H. Magnusson, Gisli Baldursson, Katrin Davidsdottir, Gyda S. Haraldsdottir, Trine Tollerup Nielsen, Esben Agerbo, Gabriel E. Hoffman, Søren Dalsgaard, Joanna Martin, Marta Ribasés, Dorret I. Boomsma, Maria Soler Artigas, Nina Roth Mota, Daniel Howrigan, Sarah E. Medland, Tetyana Zayats, Veera Manikandan, ADHD Working Group of the Psychiatric Genomics Consortium, iPSYCH-Broad Consortium, Merete Nordentoft, Ole Mors, David M. Hougaard, Preben Bo Mortensen, Mark J. Daly, Stephen V. Faraone, Hreinn Stefansson, Panos Roussos, Barbara Franke, Thomas Werge, Benjamin M. Neale, Kari Stefansson, Anders D. Børglum

**Author notes:** shared first authors (Ditte Demontis, G. Bragi Walters, Georgios Athanasiadis, Raymond Walters). Shared last authors (Barbara Franke, Thomas Werge, Benjamin M. Neale, Kari Stefansson, Anders D. Børglum). **Correspondence and requests for materials should be addressed to:** Ditte Demontis, Anders D. Børglum.

## Abstract

Attention deficit hyperactivity disorder (ADHD) is a prevalent childhood psychiatric disorder, with a major genetic component. Here we present a GWAS meta-analysis of ADHD comprising 38,691 individuals with ADHD and 186,843 controls. We identified 27 genome-wide significant loci, which is more than twice the number previously reported. Fine-mapping risk loci highlighted 76 potential risk genes enriched in genes expressed in brain, particularly the frontal cortex, and in early brain development. Overall, ADHD was associated with several brain specific neuronal sub-types and especially midbrain dopaminergic neurons. In a subsample of 17,896 exome-sequenced individuals, we identified increased load of rare protein-truncating variants in cases for a set of risk genes enriched with likely causal common variants, suggesting implication of *SORCS3* in ADHD by both common and rare variants. We found ADHD to be highly polygenic, with around seven thousand variants explaining 90% of the SNP heritability. Bivariate gaussian mixture modeling estimated that more than 90% of ADHD influencing variants are shared with other psychiatric disorders (autism, schizophrenia and depression) and phenotypes (e.g. educational attainment) when both concordant and discordant variants are considered. Additionally, we demonstrated that common variant ADHD risk was associated with impaired complex cognition such as verbal reasoning and a range of executive functions including attention.

## Introduction

Attention deficit hyperactivity disorder (ADHD) is one of the most prevalent childhood psychiatric disorders affecting around 5% of children and persists into adulthood in two-thirds of cases^1,2^. It is characterized by extensive hyperactive, impulsive and/or inattentive behaviors that impair functioning. The disorder is associated with multiple adverse outcomes such as injuries^3^, accidents^4^, depression^5^, substances use disorders^6^, aggression^7^, premature death^8^, high rate of unemployment^9^, and has large societal costs^10-12^.

ADHD has a major genetic component with an estimated twin heritability of 0.74^13^. Despite the large influence of genetics on ADHD, the complex polygenic architecture makes it difficult to unravel the underlying biological causes of the disorder. Recently, we discovered the first 12 genome-wide significant loci for ADHD^14^ in a GWAS of 20,183 cases and 35,191 controls (here referred to as ADHD2019) that combined the first wave of data from large Danish iPSYCH^15^ cohort (iPSYCH1) with 11 ADHD cohorts collected by the Psychiatric Genomics Consortium (PGC). The results implicated brain-expressed genes and demonstrated considerable genetic overlap of ADHD with a range of phenotypes, including phenotypes within psychiatric, cognitive and metabolic domains. Additionally, we established the role of common variants in ADHD, explaining around 22% of the variance in the phenotype. Besides common risk variants, analyses of whole-exome sequencing data from a subset of the iPSYCH cohort have recently shown that rare variants also contribute to the risk for ADHD^16^. The burden of rare deleterious variants in evolutionary conserved genes in ADHD cases was increased compared to controls at a level comparable to what is found in cases with autism spectrum disorder.

To better understand the biological mechanisms underlying ADHD, it is fundamental to conduct large genetic studies as demonstrated for other psychiatric disorders^17-19^. Here we present results from an updated GWAS meta-analysis of ADHD combining data from the extended Danish iPSYCH cohort (iPSYCH1 plus new iPSYCH2 data), the Icelandic deCODE cohort and the PGC, almost doubling the number of cases compared with ADHD2019. We fine-map identified risk loci and integrate our results with functional genomics data to pinpoint potential causal genes, and evaluate the burden of rare deleterious variants in top-associated genes. We characterize the polygenic architecture of ADHD and its overlap with other phenotypes by e.g. bivariate causal mixture modelling and perform polygenic score (PGS) analyses in order to test for association of ADHD-PGS with neurocognitive measures in the Philadelphia Neurodevelopmental Cohort (PNC).

## RESULTS

### Identification of new ADHD risk loci by GWAS meta-analysis

We conducted a GWAS meta-analysis based on expanded data from iPSYCH (25,895 cases; 37,148 controls), deCODE genetics (8,281 cases; 137,993 controls) and previously published data from 10 ADHD cohorts with European ancestry collected by the PGC (4,515 cases; 11,702 controls), which resulted in a total sample size in the meta-analysis of 38,691 cases and 186,843 controls (effective sample size = 51,568; cohorts are listed in Supplementary Table 1). The iPSYCH cases comprise all individuals born in Denmark between 1981 and 2008 diagnosed with ADHD up to 2016 (Ref.^20^). They were identified in the Danish Psychiatric Central Research Register^21^ and the National Patient Register^22^ based on ICD10 diagnosis codes (Online Methods). Controls are population-based individuals without a diagnosis of ADHD. deCODE cases were also clinically diagnosed according to ICD10 or identified based on medication prescribed specifically for ADHD (mostly methylphenidate). deCODE controls were not diagnosed with ADHD or other major psychiatric disorders. Ascertainment and diagnosis criteria for the PGC cohorts have been described in details previously^14^.

Quality control and imputation was done separately for each cohort (Online Methods), and GWAS results from logistic regression (using relevant covariates) in the single cohorts were combined into a GWAS meta-analysis using an inverse-variance-weighted fixed effects model^23^. The meta-analysis identified 32 lead variants (r^2^ < 0.1) located in 27 genome-wide significant loci (Figure 1; Table 1, locus plots in Supplementary Data 1), including 21 novel loci. No statistically significant heterogeneity was observed between cohorts (Supplementary Figure 1). The three strongest associated loci (*P* < 5×10^−14^) were located on chromosome 1 (in and around *PTPRF*), chromosome 5 (downstream of *MF2C*) and chromosome 11 (downstream of *METTL15*). The latter locus on chromosome 11 is a novel ADHD risk locus. Four loci on chromosomes 1, 5, 11 and 20 had secondary genome-wide significant lead variants (r^2^ < 0.1 between the index variant and the secondary lead variant within a region of 0.5 MB), but none of these remained genome-wide significant in analyses conditioning on the index variant using COJO (Supplementary Table 2).

**Figure 1.**
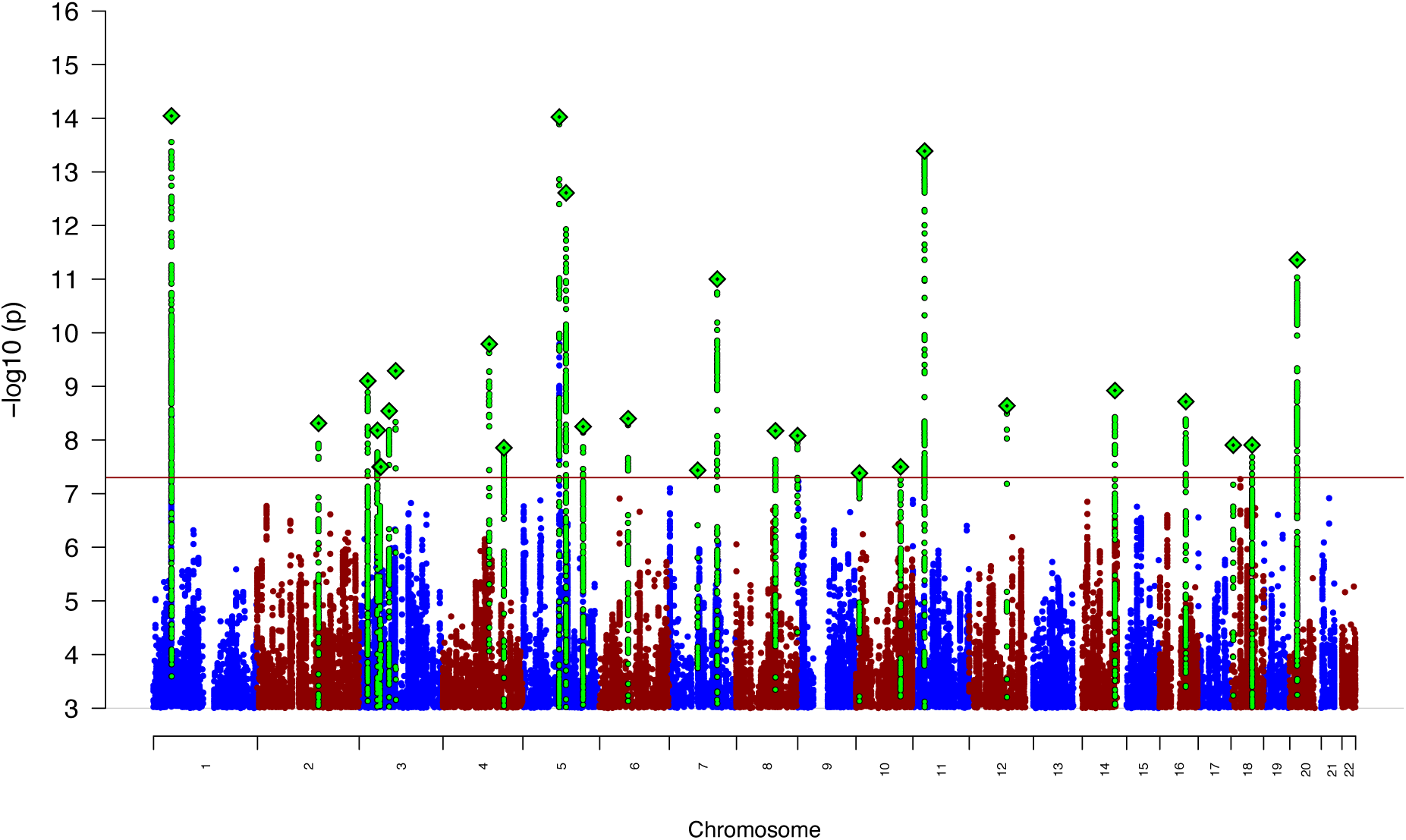
Results from GWAS meta-analysis of iPSYCH, deCODE and PGC cohorts in total including 38,899 cases and 186,843 controls. Two-sided *P*-values from meta-analysis using an inverse-variance weighted fixed effects model. Index variants in each of the genome-wide significant loci are marked as a green diamond (note that two loci on chromosome 3, index variants rs7613360 and rs2311059, are located in close proximity and therefore appears as one diamond in the plot.The red horizontal line represents the threshold for genome-wide significant association (*P* = 5×10^−8^).

**Table 1.**
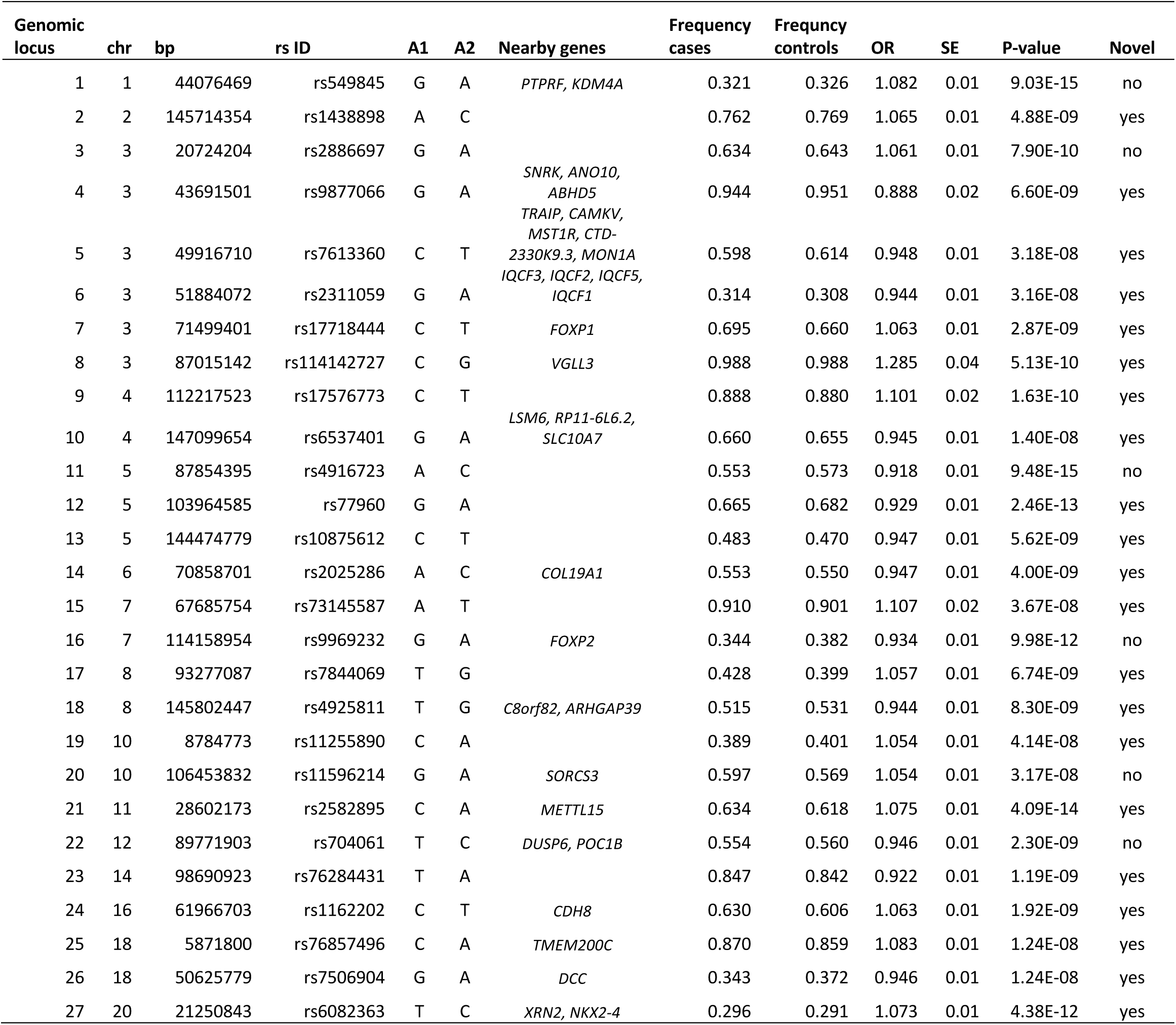
Results for the 27 genome-wide significant index variants identified in the GWAS meta-analysis of 38,691 individuals with ADHD and 186,843 controls. The location (chromosome (chr)) base position (bp) in hg19), alleles (A1 and A2), odds ratio (OR) of the effect with respect to A1, standard error (SE) and association *P*-values from inverse-variance weighted fixed effects model of the index variants are given. “Novel” indicates if the locus is a new ADHD risk locus i.e. not identified in ADHD2019 (Ref^14^). Nearby genes located within 50 kb from index variants are listed (for a list of mapped genes based on other criteria see Supplementary Table 8).

Six of the previously identified 12 loci in the ADHD2019 study^14^ were significant in the present study (Table 1), and the remaining six loci demonstrated P-values < 8×10^−4^ (Supplementary Table 3). Overall, the direction of association of the top loci (726 loci with *P* < 1×10^−4^) was consistent with the direction of association in ADHD2019 for all loci, except for one (Supplementary Table 4).

### Genetic correlations among cohorts and SNP-heritability

Genetic correlation analyses supported a high consistency in the phenotype across cohorts, including a high genetic correlation (r_g_) between iPSYCH1 and iPSYCH2 (r_g_ = 0.97; s.e. = 0.06; Supplementary Table 5). The r_g_ was 0.93 (s.e.= 0.21) between deCODE and PGC, and 0.82 (s.e. = 0.08) between iPSYCH and deCODE; none of the genetic correlations were significantly different form 1. LD score regression analysis found an intercept of 1.04 (s.e.=0.009) and ratio of 0.092 (s.e. = 0.02), the latter indicating that around 90% of the deviation from null, in the distribution of the test statistics, reflects polygenicity (QQ-plot shown in Supplementary Figure 2). The SNP heritability (h^2^_SNP_) was estimated to 0.14 (s.e. = 0.01), which is lower than the previously reported h^2^_SNP_ of 0.22^14^. The h^2^_SNP_ for iPSYCH (h^2^_SNP_ = 0.23; s.e. = 0.01) was in line with the previous finding, but lower h^2^_SNP_ was observed for PGC (h^2^_SNP_ = 0.12; s.e. = 0.03) and deCODE (h^2^_SNP_ = 0.081; s.e. = 0.014). Between-cohort heterogeneity in h^2^_SNP_ is not unusual and has been observed for other disorders like e.g. MDD^24^.

### Mapping risk variants to genes and enrichment analyses

In order to link identified risk variants to genes by incorporating functional genomics information, we first identified sets of Bayesian credible variants for each risk locus, which most likely (probability > 95%) include a causal variant (Supplementary Table 6). The sets of credible variants were linked to genes based on genomic position, and information about expression quantitative trait loci (eQTLs) and chromatin interaction mapping was derived from human brain tissue datasets implemented in FUMA^25^ (datasets selected in FUMA are listed in the Supplementary Information). Seventy-six (76) plausible ADHD risk genes were identified (Supplementary Table 7). We found that this set of genes is significantly enriched in genes upregulated during early embryonic brain development (19^th^ post-conceptual week; P=0.0002; Supplementary Figure 3) and highly enriched for genes identified in GWASs of cognition-related phenotypes and reproduction (Supplementary Figure 4). Assessment of the role of the genes in synapses were evaluated using SynGO data^26^. Nine genes mapped to SynGO annotations, and genes encoding integral components of the postsynaptic density membrane were borderline significantly enriched (5.43×10^−3^; q-value 0.022; genes *PTPRF, SORCS3, DCC*; Supplementary Figure 5; Supplementary Table 8). Additionally, enrichment of the 76 genes in biological pathways was tested using data from 26 databases, but no significant pathways showed significant enrichment after Bonferroni correction.

### Transcriptome-wide association analysis (TWAS) of the genetically regulated gene expression

To identify and prioritize ADHD risk genes we also performed a transcriptome-wide association study (TWAS) of the genetically regulated gene expression using EpiXcan^27^ and expression data from the PsychENCODE Consortium^28^ on genes as well as isoforms detected in 924 samples from the dorsolateral prefrontal cortex (DLPFC). The TWAS identified 15 genes (Supplementary Table 9) and 18 isoforms (Supplementary Table 10), which together identified 23 distinct genes (Supplementary Figure 6) with significantly different predicted gene expression in ADHD cases compared to controls (after Bonferroni correction correcting for all the 34,646 genes and isoforms tested; Supplementary Figure 6). Eight of the genes were among the 76 risk genes mapped by credible variants in FUMA. If we instead applied a less stringent correction using a false discovery rate < 5% we identified 237 genes with different predicted expression among cases and controls, of which 19 genes were also among the 76 ADHD risk. The *B4GALT2-205* isoform located in the genome-wide significant locus on chromosome 1 showed the strongest association (*P* = 7×10^−11^), with lower predicted expression in ADHD compared to controls (Supplementary Figure 7.A). The expression model for *B4GALT2-205* implicated four genome-wide significant variants. The second top gene was *PPP1R16A* (*P* = 1.4×10^−8^), which showed a predicted under-expression in cases compared to controls. The expression model for this gene implicated one genome-wide significant variant (Supplementary Figure 7.B).

### Gene-based association, tissue and cell-type specific expression of ADHD risk genes

Gene-based association analysis using MAGMA^29^ identified 45 exome-wide significant genes (*P* < 2.72×10^−6^ (0.05/18381 genes)) associated with ADHD (Supplementary Table 11). Gene association results across the entire genome were tested for a relationship with tissue specific gene expression. This showed that brain-expressed genes, and in particular genes expressed in the cortex, are associated with ADHD (Supplementary Figure 8). This result was supported by LDSC-SEG^30^ analysis, showing a significant enrichment in the heritability by variants located in genes specifically expressed in the frontal cortex (Supplementary Table 12).

Next we examined neuronal cell-type specific gene expression in ADHD using two approaches. First, we tested for enrichment of variants located in cell-specific epigenomic peaks by intersecting our genetic associations with data from two recent catalogs of the human epigenome that profile major human body cell types^31^ as well as brain-specific cell types^32^. Here we found enrichment for genes expressed in major brain neuronal cell types including both excitatory and inhibitory neurons (Supplementary Figure 9). Second, we performed cell-type specific analyses in FUMA^33^ based on single cell RNA-sequencing data. This revealed a significant association (*P* = 0.005) between ADHD-associated genes and genes expressed in dopaminergic midbrain neurons (Linnarsson midbrain data^34^; Supplementary Figure 10; Supplementary Table 13).

### Convergence of common and rare variant risk

In order to test for convergence of risk conferred by common variants and rare protein-truncating variants (rPTVs), we analyzed whole-exome sequencing data from a subset of the iPSYCH cohort consisting of 8,895 ADHD cases and 9,001 controls. We tested three gene-sets: 1) the 76 ADHD plausible risk genes identified by positional and functional annotation, 2) the 45 significant genes in the MAGMA analysis, and 3) 18 genes with at least five credible variants located in the coding region (Supplementary Table 14). While there was no indication of increased burden of rPTVs in the first gene set (*P* = 0.39), the second gene set showed borderline nominal significant enrichment (*P* = 0.05) and the set of genes identified based on credible variants was significantly enriched in cases compared to controls (*P* = 0.015). For comparison, there was no enrichment in rare synonymous variants in the latter gene-set (*P* = 0.59). When evaluating the 18 genes from the “credible gene set” individually, *SORCS3* was nominally significantly (*P* = 0.008; Supplementary Table 14) enriched in rare rPTVs in ADHD cases when compared to a combined group of iPSYCH controls and gnomAD individuals (non-psychiatric non-Finnish Europeans; N=58,121), suggesting that *SORCS3* might be implicated in ADHD both by common and rare deleterious variants (Supplementary Table 14).

### Genetic overlap of ADHD with other phenotypes

The genome-wide genetic correlation of ADHD with other phenotypes was estimated using published GWASs (258 phenotypes) and GWASs of UK Biobank data (514 phenotypes), available in LDhub^35^. ADHD was significantly correlated (*P* < 2×10^−4^) with 56 phenotypes representing domains previously found to have significant genetic correlations with ADHD: cognition (e.g. educational attainment r_g_ = -0.55, s.e. = 0.021), weight/obesity (e.g. body mass index r_g_ = 0.27, s.e. = 0.03), smoking (e.g. smoking initiation r_g_ = 0.48; s.e = 0.07), sleep (e.g. insomnia r_g_ = 0.46, s.e. = 0.05), reproduction (e.g age at first birth r_g_ = -0.65, s.e. = 0.03), and longevity (e.g. mother’s age at death r_g_ = -0.42, s.e. = 0.07). When considering other psychiatric disorders, autism spectrum disorder (ASD) (r_g_ = 0.42, s.e. = 0.05), schizophrenia (SZ) (r_g_ = 0.17, s.e. = 0.03), major depressive disorder (MDD) (r_g_ = 0.31, s.e. = 0.07), and cannabis use disorder (CUD) (rg = 0.61, s.e. = 0.04) were significantly correlated with ADHD (Supplementary Table 15). In UK Biobank data, ADHD demonstrated the strongest genetic correlation with a low overall health rating (r_g_ = 0.60, s.e. = 0.2; Supplementary Table 16).

Furthermore, we applied MiXeR^36^ which uses uni- and bivariate gaussian mixture modeling to quantify the actual number of variants that: 1) explain 90% of the SNP heritability of ADHD and 2) overlap between ADHD and other phenotypes representing domains with high genetic correlation with ADHD (psychiatric disorders, smoking behavior, weight, reproduction, and sleep were evaluated). MiXeR considers all variants irrespective of the direction of genetic correlation (i.e. both variants with same and opposite effects). Approximately 7,000 (s.e. = 285) common variants were found to influence ADHD, which is less than for SZ (8,5K; s.e. = 240), MDD (17.9K; s.e. = 1,700) and ASD (9.9K; s.e. = 1,050), and less than observed previously for bipolar disorder (BD) (8,6K, s.e. = 200)^17^. Remarkably, when considering the number of shared loci as a proportion of the total polygenicity of ADHD, the vast majority of variants influencing ADHD were also estimated to influence the other investigated psychiatric disorders (93%-94%; Figure 2; Supplementary Table 17). While the fraction of concordant variants (within the shared part) with MDD was high (96%), it was considerably lower for SZ (56%) and in-between for ASD (69%). When considering other phenotypes, insomnia demonstrated the smallest overlap with ADHD in terms of actual number of variants (4.4K, s.e. = 1,296; 62% of ADHD variants shared) while almost all variants influencing ADHD also influence educational attainment, age at first birth and smoking (Figure 2; Supplementary Table 17). For insomnia and smoking, 84% and 80% of shared variants have concordant directions, respectively, while only 22% and 19% of ADHD risk variants were concordant with educational attainment and age at first birth associated variants, respectively (Supplementary Table 17).

**Figure 2.**
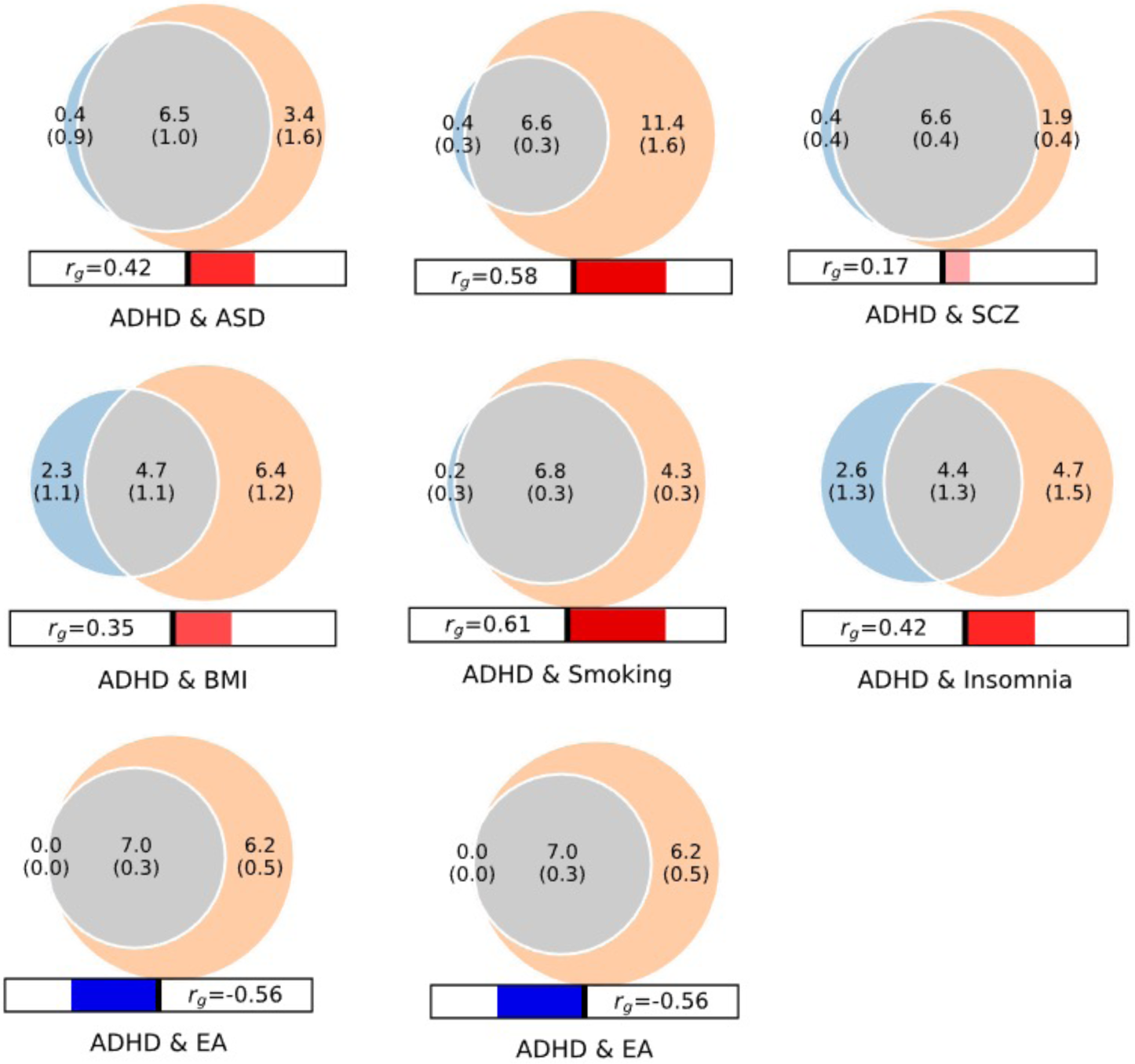
Venn diagrams showing MiXeR results of the estimated number of variants shared between ADHD and psychiatric disorders (with significant genetic correlations with ADHD) and phenotypes representing other domains with high genetic correlation with ADHD. Circles represent shared loci (gray), unique to ADHD (light blue) and unique to the phenotype of interest (orange). The number of shared variants (and standard errors) are shown in thousands. The size of the circles reflects the polygenicity of each phenotype, with larger circles corresponding to greater polygenicity. The estimated genetic correlation (*r*_g_) between ADHD and each phenotype from LDSC is shown below the corresponding Venn diagram, with an accompanying scale (−1 to +1) with blue and red representing negative and positive genetic correlations, respectively. Bivariate results for ADHD, autism spectrum disorder (ASD), major depressive disorder (MDD), schizophrenia (SCZ), body mass index (BMI), smoking initiation, insomnia, educational attainment (EA) and age at first birth are shown (see also Supplementary Table 17).

### Impact of ADHD polygenic scores on cognitive domains

In order to evaluate the impact of ADHD risk variants on cognitive domains, we assessed the association of ADHD polygenic scores (PGS) with 15 cognitive measures in the Philadelphia Neurodevelopmental Cohort (PNC)^37,38^. This cohort is from the greater Philadelphia area, and include individuals in the age of 8-21 years who received medical care at the Children’s Hospital of Philadelphia Network. The PNC cohort (v1 release) is a sample of 8,722 genotyped individuals. The subsample of 4,973 individuals with European descent was utilized in this study. The Computerized Neurocognitive Battery^39^ was used to assess cognitive performance in the subjects, the battery consists of 14 tests in 5 domains: executive-control, episodic memory, complex cognitive processing, social cognition, and sensorimotor speed. Additionally, the Wide Range Achievement Test (WRAT-4)^40^ was used as a measure to get a proxy for overall IQ^38^.

ADHD-PGS was negatively associated with seven neurocognitive domains (Figure 3) with the strongest association for the WRAT-4 test (beta = -0.09, *P* = 4.35×10^−10^). Besides that, ADHD-PGS was associated with measures of executive control (attention: beta = -0.07, *P* = 3.25×10^− 7^; working memory: beta = -0.05, *P* = 2.45×10^−3^), complex cognition (verbal reasoning: beta = -0.08, *P* = 4.74×10^−12^; non-verbal reasoning: beta = -0.06, P = 6.28×10^−4^; spatial reasoning: beta = -0.06, *P* = 5.15×10^−5^) and one measure of episodic memory (facial memory: beta = -0.05, *P* = 3.23×10^−3^)(Supplementary Table 18). The negative association of ADHD risk variants with executive functions, especially attention, is in line with the inattention problems often observed in individuals with ADHD.

**Figure 3.**
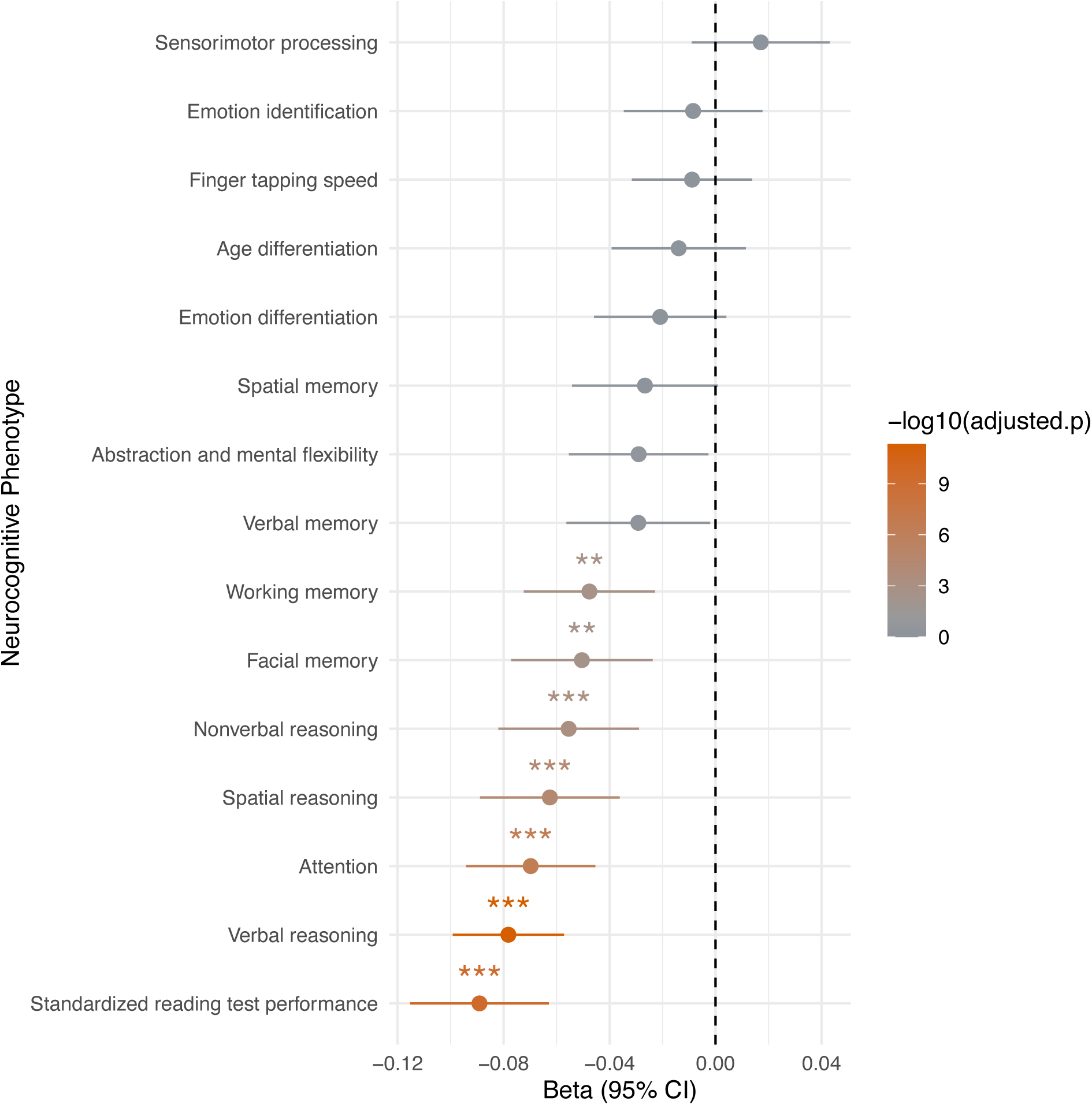
Association of ADHD-PGS with measures of cognitive abilities in the PNC cohort (N=4,973). Beta values (and standard errors indicated as horizontal bars) from linear regression testing for the association of ADHD-PGS with the 15 neurocognitive measures listed on the y-axis. The color bar at the right indicate the -log10(Bonferroni adjusted *P*-value) and significant results are indicated by stars (* *P* < 0.05; ** *P* < 0.01, *** *P* < 0.001).

## Discussion

The present study identified 27 genome-wide significant loci in the largest GWAS of ADHD to-date. We analyzed around twice as many ADHD cases as in the previous GWAS meta-analysis (ADHD2019)^14^ and more than doubled the number of associated loci, indicating that we have passed the inflection point^41^ for ADHD with respect to the rate of risk loci discovery. Six of the 12 previously identified loci were also significant in this study. Even though some previously identified loci demonstrated less association here, their associations were still strong and there was almost complete concordance in the direction of association between top-associated variants in this study and ADHD2019. The observation that previously identified loci may not reach genome-wide significance in a subsequent larger GWAS, has also been seen for other psychiatric disorders, e.g. bipolar disorder, where eight out of 19 loci were significant in a subsequent larger study^42^.

We report a lower h^2^_SNP_ for ADHD (h^2^_SNP_=0.14) than estimated previously (h^2^_SNP_=0.22). This is driven by a lower h^2^_SNP_ in the PGC and deCODE cohorts compared to iPSYCH. Different ascertainment strategies and designs among PGC cohorts could decrease the h^2^_SNP_, while lower effective sample size^43^ on Iceland, and thus fewer recent variants might bias h^2^_SNP_ downwards in the deCODE cohort^44^.

Our analyses highlighted that ADHD is a very polygenic disorder by estimating that around 7,000 common variants can explain 90% of the h^2^_SNP_. Interestingly, the estimated number of ADHD risk variants was lower than observed for three genetically correlated psychiatric disorders (SZ, MDD, ASD). It could be hypothesized that a relatively larger phenotypic/genetic heterogeneity within these three disorders (as reported in e.g. ^45,46^) could explain a part of the larger number of common risk variants influencing these phenotypes. Surprisingly, when assessing both concordant and discordant allelic directions, we found that over 90% of ADHD risk variants also influence the other investigated psychiatric disorders. This extensive sharing with ASD, SZ and MDD is at the same level as observed for SZ and bipolar disorder^36^, which are among the most genetically correlated mental disorders^47^. Notably, for MDD almost all variants shared with ADHD demonstrated concordant direction of association. The large sharing of variants influencing ADHD and other psychiatric disorders, when assessing both concordant and discordant allelic directions, suggest that the disorders are even more intermingled with respects to their common genetic architecture than previously thought based on their overall genetic correlations^36,47^. When considering common variants, the developmental trajectory towards ADHD might therefore be influenced by variants involved in several psychiatric disorders but with disorder-specific effect sizes rather than actual ADHD-specific risk variants. We also note, that all variants that influence ADHD overlap with educational attainment^48^ and that around 22% of ADHD risk variants are associated with increased educational attainment while the vast majority (78%) are associated with decreased educational attainment. This is consistent with the overall negative genetic correlation with educational attainment and the positive genetic correlation of ADHD with e.g. ASD that has a positive genetic correlation with educational attainment^45,49^. Thus, shared ADHD/ASD variants might be a part of the 22% of variants concordant between ADHD and educational attainment. Likewise, risk variants that differentiate ADHD from ASD might primarily be found among those that overlap with decreased educational attainment, which is consistent with results recently been reported in ADHD-ASD differentiating genetic analyses^50^.

Fine-mapping of the 27 loci identified credible variants that in general had low posterior probabilities. Only four variants had posterior probabilities greater than 0.5 in all three fine-mapping methods, and none were linked to specific genes based on our functional annotation analyses. Linking the credible variants to genes by integration with functional genomics data identified 76 likely risk genes, which were enriched among genes upregulated during early embryonic development and involved in cognitive abilities identified by GWAS of cognitive phenotypes. Among these genes were *PPP1R16A* and *B4GALT2* (mapped by psychENCODE eQTLs; Supplementary Figure 11.A and B), which were also the top-ranking genes in our TWAS of DLPFC expression, both showing a predicted decreased expression in cases compared to controls. These genes have not previously been linked to psychiatric disorders, but both have been linked to educational attainment^48^ with increased predicted expression in cortical brain regions associated with increased measures of cognitive ability (assessed in gene2pheno/PhenomeXcan_SingleTissue^51^, *PPP1R16A* (cortex): educational years^48^ *P* = 2.7×10^−6^; UK Biobank college or university degree P = 1.05×10^−7^; *B4GALT2* (frontal cortex) *P=*1.98×10^−7^). The set of risk genes also included *PTPRF, SORCS3* and *DCC* which encode integral components of the postsynaptic density membrane. Involvement of postsynaptic components in the pathology of ADHD has been reported previously (identified by MTAG analyses of ADHD and related psychiatric disorders^52^) and also for other psychiatric disorders^53^. We would also like to highlight *FOXP1* and *FOXP2*, which were among the 76 risk genes. The genome-wide significant signal was located within the transcribed regions of both genes, and were additionally implicated in ADHD by genome-wide significant variants being eQTLs (*FOXP2*, Supplementary Figure 11.C) or located in chromatin interacting regions (*FOXP1*, Supplementary Figure 11.D) in brain tissue. *FOXP2* was identified in the ADHD2019 study^14^, and the association was even stronger in the current study. This gene was also recently identified as a risk gene for cannabis use disorder^54^. *FOXP1* is a new ADHD risk gene, which has previously been associated with schizophrenia^19^. Both *FOXP1* and *FOXP2* encode transcription factors that can heterodimerize to regulate transcription in brain tissues^55,56^. Highly penetrant rare variants have implicated the genes in language and speech disorders and intellectual disability^57^. The relative levels and the specific combination of *FOXP1* and *FOXP2* hetero/homo-dimers seem to regulate transcription of target genes differently^58^, and thus it could be speculated that deviations in the relative levels of the proteins (altered by the regulatory common risk variants) might affect brain development.

We report convergence of common and rare variants in a set of 18 genes defined by location of credible variants. Thirteen of the genes were hit by rPTVs and for eight of them there was a higher load in cases compared to controls. The signal was not driven by a few genes but by several genes with an increased burden of rPTVs. Notably, in particular *SORCS3* seems to be implicated in ADHD by both common and rare variants. Common variants in *SORCS3* show strong pleiotropic effects across several major psychiatric disorders^47^, but to our knowledge, rare variant analyses have not implicated *SORCS3* in psychiatric disorders before. Our results add to the emerging picture of overlap between genes and pathways affected by common and rare variants in psychiatric disorders^53,59, 60,61^.

We found that ADHD risk was associated with common variants located in genes significantly expressed in the brain, especially the frontal cortex. We also observed an enrichment of ADHD risk variants in genes expressed in major cell types of the brain including both excitatory and inhibitory neurons and in midbrain dopaminergic neurons. The findings for frontal cortex and dopamine neurons fit well with the motor, reward and executive function deficits associated with ADHD; the frontal cortex is involved in executive functions e.g. attention and working memory^62^, and midbrain dopaminergic neurons are essential for controlling key functions, such as voluntary movement^63^ and reward processing^64^. This interpretation is further supported by our ADHD-PGS analyses in PNC which revealed that common ADHD risk variants impair several domains of cognitive abilities, including attention and working memory.

The PGS analyses in PNC identified strong association of polygenic ADHD risk with decreased overall IQ (approximated by the WRAT test scores), which is in line with the high negative genetic correlation of ADHD with educational attainment and the observation that 78% of all ADHD risk variants are associated with decreased educational attainment. Interestingly, we found that ADHD-PGS associates with decreased attention, which is a key ADHD symptom, and with impairments in measures of other cognitive traits such as working memory. Smaller studies have previously analyzed the impact of ADHD-PGS on executive functions with mixed results^65-68^. This study robustly identifies specific cognitive domains impacted by ADHD-PGS and our results support ADHD-PGS to be negatively associated with neurocognitive performance.

In summary, we identified new ADHD risk loci, highlighted candidate causal genes and implicated genes expressed in frontal cortex and several brain specific neuronal sub-types in ADHD. Our analyses revealed ADHD to be highly polygenic, influenced by thousands of variants, of which the vast majority also influence other psychiatric disorders with concordant or discordant effects. Additionally, we demonstrated that common variant ADHD risk has an impairing impact on a range of executive functions. Overall, the results advance our understanding of the underlying biology of ADHD, and reveal novel aspects of ADHD’s polygenic architecture, its relationship with other phenotypes and its impact on cognitive domains.

## METHODS

### Samples, quality control and imputation

#### iPSYCH

The iPSYCH^15,20^ cohort consists of 129,950 genotyped individuals, among which 85,891 are cases diagnosed with at least one of six mental disorders (i.e. ADHD, SZ, BD, MDD, ASD, post-partum disorder) and the remaining are population-based controls. Samples were selected from a baseline birth cohort comprising all singletons born in Denmark between May 1, 1981, and December 31, 2008, who were residents in Denmark on their first birthday and who have a known mother (N = 1,657,449). ADHD cases were diagnosed by psychiatrists at in- or out-patient clinics according to the ICD10 criteria (F90.0, F90.1, F98.8 diagnosis codes) identified using the Danish Psychiatric Central Research Register^21^ and the Danish National Patient register^22^. Diagnoses were given in 2016 or earlier for individuals at least 1 year old. Controls were randomly selected from the same nationwide birth cohort and not diagnosed with ADHD. The study was approved by the Danish Data Protection Agency and the Scientific Ethics Committee in Denmark.

The samples were genotyped in two genotyping rounds referred to as iPSYCH1 and iPSYCH2. DNA extraction and subsequent whole-genome amplification was performed as previously described^15^. iPSYCH1 samples were genotyped using Illumina’s PsychChip array and iPSYCH2 samples using the Illumina’s global screening array v.2 (Illumina, CA, San Diego, USA). iPSYCH1 genotypes were called using GenCall and Birdseed and iPSYCH genotypes were called using GenTrain V3.

Pre-imputation quality control and imputation was performed on genotypes from the full set of genotyped individuals for iPSYCH1 and iPSYCH2 separately. Quality control, imputation and primary association analyses were done using the bioinformatics pipeline “Ricopili”^69^. Subjects and variants were included in the imputation based on the following quality control parameters: variant call rate > 0.95 (before sample removal), subject call rate > 0.95, autosomal heterozygosity deviation (| Fhet | < 0.2), variant call rate > 0.98 (after sample removal), difference in variant missingness between cases and controls < 0.02, and SNP Hardy-Weinberg equilibrium (HWE) (P > 10^−6^ in controls or P > 10^−10^ in cases). The iPSYCH1 samples were genotyped in 23 genotyping waves and thus additional steps were taken in order to eliminate potential batch effects. Only variants present in more than 20 waves and with no significant association with wave status, were retained. Imputation was done using the prephasing/imputation stepwise approach implemented in EAGLE v2.3.5^70^ and Minimac^71^, using the Haplotype Reference Consortium^72^ panel v1.0. iPSYCH1 comprised imputed genotypes from 20,175 ADHD cases and 25,836 population-based controls without ADHD, and iPSYCH2 contained imputed genotypes on 10,624 ADHD cases and 18,255 controls.

Best guest genotypes from iPSYCH1 and iPSYCH2 were merged in order to identify potential duplicated samples and related individuals within and across the entire sample. Related (duplicated samples) were identified by “identity by state” analysis in plink v1.9, and one individual was excluded from pairs of subjects with pi_hat > 0.2. For this a set pruned best guess genotypes (imputation INFO score > 0.8; r^2^ < 0.075; markers located in long range LD regions defined by Price et al.^73^ excluded) with minor allele frequency > 0.05 and no deviation from Hardy-Weinberg equilibrium (HWE P > 1× 10^−4^) was used. This step removed 5,326 individuals.

Genetic outliers were identified by principal component analysis (PCA) which was performed separately for iPSYCH1 and iPSYCH2 using high quality variants as described above and the software Eigensoft^74^. Non-European individuals were excluded if their principal component (PC) values for PC1 and PC2 were greater than six standard deviations from the centre of an ellipsoid where the centre was based on the mean values of PC1 and PC2 of a sub-sample of Danish individuals. The subsample of Danes was defined using register information about birth country of the individuals and their parents which were required to be Denmark. After exclusion of non-European samples PCAs were re-run to exclude remaining population stratification, which was done by visual inspection of PCA plots. After QC the iPSYCH1 ADHD sample included 38,899 individuals and iPSYCH2 included 24,144 individuals.

#### deCODE

The deCODE cohort consisted of ADHD cases with a clinical diagnosis of ADHD according to the ICD10 criteria (ICD10-F90, F90.1, F98.8) or individuals that have been prescribed medication specific for ADHD symptoms (ATC-NA06BA, mostly methylphenidate). The control sample did not contain individuals with a diagnosis of SZ, BD, ASD or self-reported ADHD symptoms or diagnosis. The study was approved by the National Bioethics Committee of Iceland (VSN 15-047) and all participants who donated samples gave informed consent. Samples were assayed with several Illumina arrays at deCODE genetics and genotypes called using GraphTyper2^75^. SNPs with low call rate (<95%), significant deviation from Hardy-Weinberg equilibrium (P<0.001), and excessive inheritance error rates (>0.001) were excluded. Variant imputation was performed based on the IMPUTE HMM model and long-range phasing, as described previously^76^.

#### PGC cohorts

We used summary statistics from the 10 PGC cohorts with European ancestry generated as a part of our previous GWAS meta-analysis of ADHD. Detailed information about cohort design, genotyping, QC and imputation can be found in Demontis and Walters et al^14^.

### GWAS meta-analysis of ADHD

GWASs were performed separately for iPSYCH1 (17,019 cases and 21,880 controls) and iPSYCH2 (8,876 cases and 15,268 controls) using dosages for imputed genotypes and additive logistic regression with the first 10 PCs (from the final PCAs) as covariates using PLINK v1.9. GWAS of deCODE samples (8,281 ADHD cases; 137,993 controls) was done using dosage data and logistic regression with sex, age, and county of origin as covariates. To account for inflation due to population stratification and cryptic relatedness, test statistics were divided by an inflation factor (lambda= 1.23) estimated from LD score regression as done previously^54^. Findings from analyses of the genetic structure of the Icelandic population by Price et al.^77^ support that lambda correction will ensure proper correction without false positives. Subsequently alleles were converted to match HRC alleles.

For the PGC cohorts we used GWAS summary statistics for each of the 10 European PGC cohorts generated as a part of our previous GWAS meta-analysis^14^.

Summary statistics from GWAS of the individual cohorts, containing variants with imputation quality (INFO score) > 0.8 and minor allele frequency > 0.01, were meta-analyzed using an fixed effects standard error weighted meta-analysis using METAL (version 2011-03-25)^23^. Only variants supported by an effective sample size greater than 60% were retained in the final summary statistics (6,774,228 variants).

Concordance in the direction of associations with associations in the ADHD2019 data were evaluated by a sign-test at different p-value thresholds in the present GWAS (see thresholds in Supplementary Table 4).

### Conditional analysis

We identified potential independent genome-wide significant lead variants for four loci located on chromosome 1 (two secondary lead variants), 5, 11 and 20. In order to evaluate if these variants were secondary independent lead variants, we performed association analyses of the secondary lead variants while conditioning of the index variant in the locus using COJO as implemented in GCTA^78^.

### Identification of sets of credible variants

In order to identify sets of causal variants we fine-mapped each of the 27 genome-wide loci using three fine-mapping tools, FINEMAP v. 1.3.1(Ref. ^79^), PAINTOR v.3.0 (Ref.^80^) and CAVIARBF v.0.2.1 (Ref.^81^), using CAUSALdb-finemapping-pip downloaded from https://github.com/mulinlab/CAUSALdb-finemapping-pip^82^. Since no secondary lead variants remained genome-wide significant after conditional analyses, one causal variant was assumed per locus. Variants located in a region of 1MB around index variants were included in the analyses. We used a threshold 95% for the total posterior probability of the variants included in the credible sets and only variants claimed to be within the set by all three methods were included in the final credible set for each locus.

### Genetic correlations among ADHD cohorts and SNP heritability

SNP heritability (h^2^_SNP_) and pair-wise genetic correlation among the cohorts were calculated using LD score regression^83^ analysis of summary statistics from GWAS of deCODE samples, meta-analysis of iPSYCH1+iPSYCH2 and meta-analysis of the 10 PGC cohorts (applying the same approach as described for the meta-analysis of all cohorts). Conversion of h^2^_SNP_ estimates from observed scale to the liability scale was done using a population prevalence of 5%. Test for significant differences in h^2^_SNP_ between cohorts was done using a Z-test.

### Mapping of risk genes, enrichment and pathway analyses

To link identified risk variants to genes, we used the set of credible variants (identified as described above) for each locus and linked variants to genes based on genomic position and functional annotations in FUMA^25^. Protein coding genes were mapped if they were located with a distance of 10Kb up- or downstream index variants or if a credible variant was annotated to the gene based on eQTL data or chromatin interaction data from human brain (data sets used in the mapping can be found in the Supplementary Note). No additional variant filtering by functional annotation was applied in the eQTL and chromatin interaction mapping. The mapping linked credible variants to 76 ADHD risk genes. These genes were used in gene-set enrichment analyses in the GENE2FUNC module in FUMA. Enrichment of ADHD risk genes among predefined sets of differently expressed genes in GTEx (54 tissue types) and Brainspan (29 different ages of samples and 11 general developmental stages) data using hypergeometric test and protein coding genes were chosen as background genes.

SynGO^26^ (dataset version: 20210225) was used to test for enrichment among the 76 risk genes for genes involved in synaptic processes and locations. We analysed for enrichment in two subsets; “biological process” (201 gene sets) and “cellular component” (92 gene sets). We controlled using a background set of “brain expressed” genes provided by the SynGo platform (defined as ‘expressed in any GTEx v7 brain tissues) containing 18,035 unique genes of which 1,225 overlap with SynGO annotated genes. For each ontology term, a one-sided Fisher exact test was performed to compare the list of ADHD risk genes and the selected background set. To find enriched terms within the entire SynGO ontology, the most specific term is selected where each ‘gene cluster’ (unique set of genes) is found and then multiple testing correction is applied using False Discovery Rate (FDR) on the subset of terms that contain these ‘gene clusters’. Only ontology terms with gene sets with a minimum of three gene were included in the enrichment analysis.

The set of 76 mapped ADHD risk genes were also used to test for enrichment in pathways/gene sets using Enrichr^84,85^ and its implemented databses (26 databases). Only pathways enriched with more than two genes were considered. We made a conservative approach and only considered pathways to be significant if the within database adjusted P-value was smaller than 0.002 (0.05/26 databases evaluated). After correction for the number of data bases no significantly enriched pathways were identified. Additionally, we tested for enrichment among the 76 genes of genes reported from the GWAS catalog (2019) and UK biobank GWASs (v1), and used https://appyters.maayanlab.cloud/Enrichr_Manhattan_Plot/ to visualize the results.

### Transcriptomic imputation model construction and TWAS

Transcriptomic imputation models were constructed as previously described^27^ for dorso-lateral prefrontal cortex (DLPFC) transcript levels^86^. The genetic dataset of the PsychENCODE cohort was uniformly processed for quality control (QC) steps before genotype imputation. The analysis was restricted to samples with European ancestry as previously described^27^. Genotypes were imputed using the University of Michigan server^87^ with the Haplotype Reference Consortium (HRC) reference panel^88^. Gene expression information (both at the level of gene and transcript) was derived from RNA-seq counts which were adjusted for known and hidden confounds, followed by quantile normalization^86^. For the construction of the transcriptomic imputation models we used EpiXcan^27^, an elastic net based method, which weighs SNPs based on available epigenetic annotation information^89^. We performed the transcript-trait association analysis for ADHD as previously described^27^. Briefly, we applied the S-PrediXcan method^27^ to integrate the ADHD GWAS meta-analysis summary statistics and the transcriptomic imputation models constructed above to obtain association results at both the level of genes and transcripts.

### Gene-based association and tissue-specific expression of ADHD risk genes

We used MAGMA v1.08 implemented in FUMA v1.3.6a (Ref.^25^) to perform gene-based association analysis using the full summary statistics from the GWAS meta-analysis. Genome-wide significance was assessed through Bonferroni correction for the number of genes tested (*P* = 0.05/18381 = 2.72×10^−6^).

Th relationships between tissue specific gene expression profiles and ADHD-gene associations was tested using MAGMA gene-property analysis of expression data from GTEx (54 tissue types) and BrainSpan (29 brain samples at different age) available in FUMA (See Supplementary Information for data sets selected in FUMA).

Enrichment in h^2^_SNP_ of ADHD associated variants located in or close to genes expressed in specific brain regions was estimated using LDSC-SEG^30^. Annotations indicating specific expression in 13 brain regions from the GTEx gene-expression database was downloaded from: https://alkesgroup.broadinstitute.org/LDSCORE/LDSC_SEG_ldscores/.

### Cell type-specific expression of ADHD risk genes

We tested for enrichment in the ADHD h^2^_SNP_ heritability of variants located in cell type specific epigenetic peaks by examining the overlap of common genetic risk variants with open chromatin from a DHS study (DNase I hypersensitive sites) profiling major human cell types^31^ and an scATAC-seq study (single-cell assay for transposase accessible chromatin)^32^ using LD-score partitioned heritability approach^90^. All regions of open chromatin were extended by 500 base pairs in either direction. The broad MHC-region (hg19 chr6:25-35MB) was excluded due to its extensive and complex LD structure, but otherwise default parameters were used for the algorithm.

Additionally, we preformed cell-type specific analyses implemented in FUMA, using data from 13 single-cell RNA sequencing data sets from human brain (data sets listed in the Supplementary Information). The method is described in details in Watanabe et al.^33^. In short, the method uses MAGMA gene-property analysis to test for association between cell specific gene expression and ADHD-gene association and correction for multiple testing corrects for all tested cell types across datasets. In a second step systematical step-wise conditional analysis per dataset is performed in order to correct for false positives when there is high correlation in expression profiles among cell-types, only cell-type specific expression of ADHD risk genes in DA1 neurons remained significant after this step.

### Overlap of common ADHD risk variants with rPTVs

We analyzed the overlap of common variants with rPTVs in a subset of iPSYCH samples that has also been whole exome-sequenced. DNA was extracted from dried blood spot samples of the study subjects and whole genome amplified in triplicates^91,92^, the coding regions of the genome were extracted using the llumina Nextera capture kit and sequencing was performed in multiple waves (Pilot 1, Wave 1, Wave 2 and Wave 3) using the Illumina HiSeq platform at the Broad Institute.

A major part of the data (Pilot 1, Wave 1, Wave 2) were also included in the recent study by Satterstrom et al.^16^, and the same quality control procedure was applied here. In short, the sequencing data were aligned to the reference genome using the BWA^93^ (Hg19) and genotype calling was done using the best practice recommended by the Genome Analysis Toolkit^94^ (GATK) v.3.4, and additional QC steps performed using Hail (Hail Team. Hail 0.2. https://github.com/hail-is/hail). All variants annotated to ACMG^95^ genes were removed due to Danish regulations. Samples were removed if they lacked complete phenotype information, inconsistencies of the imputed sex with the reported sex, if they were duplicates or being genetic outliers identified by principal component analysis (using a set of common variants and the software Eigensoft^74^), if they had an estimated level of contamination > 5% or if they had an estimated level of chimeric reads > 5%.

Only autosomal genotypes were included in our analyses. Genotypes were removed if they did not pass GATK variant quality score recalibration (VQSR) and had read depths < 10 or > 1,000. Homozygous alleles were removed if they had reference calls with genotype quality < 25, homozygous alternate alleles with PL(HomRef) less than 25 or < 90% reads supporting alternate allele. Heterozygote alleles were removed if they had PL(HomRef) < 25 or < 25% reads supporting the alternate allele, less than < 90% informative reads, or a probability of the allele balance calculated from a binomial distribution centered on 0.5 less than 1×10^−9^. After these genotype filters, variants with a call rate < 90% were removed, then samples with a call rate < 95% were removed and then additional removal of variants with a call rate < 95%. Additionally, one of each pair of related samples was removed, from pairs with pi-hat values ≥0.2. After QC, the number of individuals were 8,895 ADHD cases and 9,001 controls.

The QCed variants were annotated using SnpEff^96^ version 4.3t. The variants were also annotated with information about allele counts in the gnomAD^97^ exomes r2.1.1 database using SnpSift^96^ version 4.3t. Variants were only included if they were located in consensus high-confident regions with high read depth in both iPSYCH and gnomAD (80% of the samples in both datasets had at least 10× sequencing coverage in the region). Variants were defined as rPTVs if they were annotated as having large effects on gene function (nonsense variant, frameshift, splice site). We defined a variant as being rare if it had an allele count of five or less across the combination of the full iPSYCH exome-sequencing dataset (n=28,448) and non-Finnish Europeans in the nonpsychiatric gnomAD exome database (n = 44,779).

We for increased burden of rPTVs in ADHD compared to controls in three gene-sets (1) the 76 genes linked to credible variants based on position and functional genomic data 2) the 45 exome-wide significant genes identified in MAGMA analysis (3) Genes with at least five credible variants within the coding regions. The requirement of five credible variants was chosen in order to prioritize the most likely causal genes. This threshold excluded eight genes located in the same locus covering a broad LD region on chromosome 3 (Supplementary Data 1; page 25). Additionally, two other genes with less than five credible variants were excluded located in two other loci on chromosome 3.

The burden of rPTVs and rare synonymous (rSYNs) in cases compared to controls was tested for the three gene-sets with logistic regression corrected using the following covariates: birth year, sex, first ten principal components, number of rSYN, percentage of target with coverage > 20x, mean read depth at sites within the exome target passing VQSR, total number of variants, sequencing wave.

Only significant enrichment in the set 18 genes identified based on credible variants was found. We therefore looked specifically into these genes in order to identify if the signal was driven by specific genes. rPTVs were found in 13 of the genes and out of these eight genes had more rPTVs in cases compared to controls when looking at the raw-counts (Supplementary Table 14). We performed gene-based burden test using EPACTS (https://genome.sph.umich.edu/wiki/EPACTS) and logistic Wald test (correcting using the covariates as described above). Additionally, in order to increase power to detect increased burden of rPTVs at the gene-level in ADHD cases, we combined iPSYCH controls with information about rPTVs in gnomAD (non-Finish European individuals), done as described previously^16^. We performed gene-based test using Fisher’s exact test and only genes with higher number of rPTVs in cases compared to controls in the iPSYCH data were considered.

### Genetic overlap with other phenotypes

We estimated genetic correlations of ADHD with other phenotypes in LDhub^35^ (published GWASs: 255 phenotypes; UK Biobank GWASs: 514 phenotypes). Additionally, genetic correlations with three phenotypes not available in LDhub (cannabis use disorder^54^, smoking initiation^98^ and education attainment^48^) were estimated locally using LD score regression^83^.

We applied MiXeR^36^ to our ADHD GWAS summary statistics and a selection of complex traits (Supplementary Table 17) to quantify (i) the number of variants influencing each trait and (ii) the genetic overlap between ADHD and each of the other traits. We used MiXeR with default settings (https://github.com/precimed/mixer) in a two-step process: 1) We ran a univariate model for each trait to produce estimates of “polygenicity” (i.e. the proportion of non-null SNPs) and “discoverability’” (i.e. the variance of effect sizes of non-null SNPs). 2) We then used the variance estimates from the previous univariate step to run the bivariate model in a pairwise fashion (i.e. ADHD vs. each of the other traits) which produced estimates of four components representing (i) null SNPs in both traits; (ii) SNPs with a specific effect on the first or on the second trait; and (iii) SNPs with a nonzero effect on both traits. These four parameters can be mathematically combined in the final estimates of a polygenic overlap and genetic correlation (for details on the method see also^17^). The estimates reported explain 90% of h^2^_SNP_.

### PGS analysis of cognitive measures in PNC

PGS analysis was performed on 4,973 individuals of European ancestry from the Philadelphia Neurodevelopmental Cohort (PNC), ages 8-21. Genotypes used for PGS generation were from the first PNC release (dbGaP phs000607.v1.p1), while the neurocognitive phenotypes used were from the third release (dbGaP phs000607.v3.p2). Pre-imputation quality control steps included removing individuals whose genotypically inferred and phenotypically reported sex did not align, those with heterozygosity rates ± three standard deviations from the mean, those who did not meet the individual-level missingness filter of 0.05, and those who did not meet the identity by descent (IBD) filter (PI-HAT > 0.185). Additionally, SNPs were removed if they had high missingness rate (> 0.05), high deviation from Hardy-Weinberg equilibrium (HWE) (*P* < 0.00001), and low minor allele frequency (MAF) filters (MAF < 0.01). Genotype imputation was performed on the Michigan Imputation Server (https://imputationserver.sph.umich.edu/index.html#!), using the reference panel HRC r1.1 2016 and selecting the “Mixed population” option. Post-imputation processing included removing SNPs with an imputation R^2^ < 0.03 and filtering based on the HWE, MAF, and missingness thresholds stated above. Outlier individuals were identified and removed by plotting the first two Multi-Dimensional Scaling (MDS) dimensions. Individuals of European ancestry were identified using the GemTools package (http://www.compgen.pitt.edu/GemTools/GEM%20Documentation.pdf) and Ward’s hierarchical clustering in R.

The software PRS-CS^99^ was used to process ADHD GWAS summary statistics and assign per-allele posterior SNP effect sizes. A European LD reference panel generated from the 1000 Genomes Project data (can be downloaded here: https://github.com/getian107/PRScs) was utilized. The following default settings were used for PRS-CS: parameter a in the γ-γ prior = 1, parameter b in the γ-γ prior = 0.5, MCMC iterations = 1000, number of burn-in iterations = 500, and thinning of the Markov chain factor = 5. Additionally, the global shrinkage parameter phi was determined using a fully Bayesian method. Plink v2.0^100^ was then used to calculate individual-level ADHD PGS. Linear regression was used to test the association between ADHD PGS and neurocognitive neurocognitive phenotypes measured in the PNC. The neurocognitive measures were obtained using the Computerized Neurocognitive Battery, which consists of 14 tests in 5 domains: executive-control, episodic memory, complex cognitive processing, social cognition, and sensorimotor speed. The battery has been is described in details elsewhere (Ref^39^). Additionally, association of ADHD-PGS with results from the Wide Range Achievement Test (WRAT-4)^40^ was analyzed.

Neurocognitive phenotype metrics and the ADHD-PGS values were each scaled such that the mean value is 0 and the standard deviation is 1 (using the function scale() in base R). Age (at time of neurocognitive testing), age squared, genotyping batch, sex, and the first 10 MDS dimensions were used as covariates. The total variance explained by ADHD-PGS and model covariates for each neurocognitive phenotype was reported using Adjusted R^2^. Additionally, the variance explained by ADHD-PGS and each covariate individually was calculated in R using a variance partitioning tool (https://github.com/GabrielHoffman/misc_vp/blob/master/calcVarPart.R). Reported *P*-values were Bonferroni adjusted to account for the number of independent tests performed.

## Data Availability

All data produced in the present study are available upon reasonable request to the authors

## Acknowledgements

D.D. was supported by the Novo Nordisk Foundation (NNF20OC0065561), the Lundbeck Foundation (R344-2020-1060) the European Union’s Horizon 2020 research and innovation programme under grant agreement No. 965381(TIMESPAN).

Research reported in this publication was supported by the National Institute Of Mental Health of the National Institutes of Health under Award Number R01MH124851. The content is solely the responsibility of the authors and does not necessarily represent the official views of the National Institutes of Health. The work was also supported by the European College of Neuropsychopharmacology (ECNP) Network “ADHD Across the Lifespan”. B.F. was also supported by funding from the European Community’s Horizon 2020 Programme (H2020/2014 – 2020) under grant agreements No. 728018 (Eat2beNICE) and No. 847879 (PRIME). B.F. also received relevant funding from the Netherlands Organization for Scientific Research (NWO) for the Dutch National Science Agenda NeurolabNL project (grant 400-17-602).

S.E.M. was funded by NHMRC grants APP1172917, APP1158125 and APP1103623.

This work was supported by the Instituto de Salud Carlos III (PI19/01224, PI20/0004), by the Pla Estratègic de Recerca i Innovació en Salut (PERIS), Generalitat de Catalunya (METAL-Cat; SLT006/17/287); by the Agència de Gestió d’Ajuts Universitaris i de Recerca AGAUR, Generalitat de Catalunya (2017SGR1461), Ministry of Science, Innovation and Universities (IJC2018-035346-I to M.S.A); by the European Regional Development Fund (ERDF) and by “la Marató de TV3” (092330/31) and the ECNP Network ‘ADHD across the Lifespan’ (https://www.ecnp.eu/researchinnovation/ECNP-networks/List-ECNP-Networks/).

T.Z. is funded by NIH, Grant No. R37MH107649-07S1 and by Research Council of Norway, NRC, Grant No. 288083.

This study was also supported by the National Institutes of Health (NIH), Bethesda, MD under award numbers T32MH087004 (to K.T.), K08MH122911 (to G.V.), R01MH125246 (to P.R.) and U01MH116442 (to P.R.).

## Author contributions

**Analysis:** D.D., G.B.W., G.A., R.W., K.T., L.F., G.V., J.B., B.Z., W.Z., J.D., S.H.M. and T.T.N. **Sample and/or data provider and processing:** D.D, G.B.W, J.G., T.D.A., J.D., F.K.S, J.B.G., M.B.H., O.O.G., G.B., K.D., G.S.H., ADHD Working Group of the Psychiatric Genomics Consortium (PGC), iPSYCH-Broad Consortium, E.A., G.E.H., M.N., O.M., D.M.H., P.B.M., M.J.D., H.S., T.W., B.M.N., K.S. and A.D.B. **Writing:** D.D., G.B.W., K.T., G.A., G.V., J.B., H.S. and A.D.B. **Core revision:** D.D., G.B.W., G.A., R.W., K.T., G.V., J.B., S.D., J.M., M.R., F.K.S, D.I.B., M.S.A., N.R.M.,D.H., S.E.M., T.Z., V.M., S.V.F., H.S., P.R., B.F., B.M.N., K.S. and A.D.B. **Study direction:** D.D. andA.D.B. **Study supervision:** D.D., G.B.W., H.S., P.R., B.F., T.W., B.M.N., K.S. and A.D.B. All authors contributed with critical revision of the manuscript.

## Data availability

Summary statistics from the ADHD GWAS meta-analysis is available for download at the PGC website (https://www.med.unc.edu/pgc/download-results/).

## Code availability

No previously unreported custom computer code or algorithm were used to generate results.

