## Supplementary Information and Figures for "Genome-wide analyses of ADHD identify 27 risk loci, refine the genetic architecture and implicate several cognitive domains"

#### eQTL datasets in FUMA used for gene-mapping

BrainSeq_ge_brain.txt.gz
PsychENCODE/PsychENCODE_eQTLs.txt.gz
CMC/CMC_SVA_cis.txt.gz
CMC/CMC_SVA_trans.txt.gz
CMC/CMC_NoSVA_cis.txt.gz
CMC/CMC_NoSVA_trans.txt.gz
BRAINEAC/CRBL.txt.gz
BRAINEAC/FCTX.txt.gz
BRAINEAC/HIPP.txt.gz
BRAINEAC/MEDU.txt.gz
BRAINEAC/OCTX.txt.gz
BRAINEAC/PUTM.txt.gz
BRAINEAC/SNIG.txt.gz
BRAINEAC/TCTX.txt.gz
BRAINEAC/THAL.txt.gz
BRAINEAC/WHMT.txt.gz
BRAINEAC/aveALL.txt.gz
GTEx/v8/Brain_Amygdala.txt.gz
GTEx/v8/Brain_Anterior_cingulate_cortex_BA24.txt.gz
GTEx/v8/Brain_Caudate_basal_ganglia.txt.gz
GTEx/v8/Brain_Cerebellar_Hemisphere.txt.gz
GTEx/v8/Brain_Cerebellum.txt.gz
GTEx/v8/Brain_Cortex.txt.gz
GTEx/v8/Brain_Frontal_Cortex_BA9.txt.gz
GTEx/v8/Brain_Hippocampus.txt.gz
GTEx/v8/Brain_Hypothalamus.txt.gz
GTEx/v8/Brain_Nucleus_accumbens_basal_ganglia.txt.gz
GTEx/v8/Brain_Putamen_basal_ganglia.txt.gz
GTEx/v8/Brain_Spinal_cord_cervical_c-1.txt.gz
GTEx/v8/Brain_Substantia_nigra.txt.gz

#### Chromatin interaction datasets in FUMA used for gene mapping

EP/PsychENCODE/EP_links_oneway.txt.gz:
HiC/PsychENCODE/Promoter_anchored_loops.txt.gz:
HiC/Giusti-Rodriguez_et_al_2019/Adult_Cortex.txt.gz:
HiC/Giusti-Rodriguez_et_al_2019/Fetal_Cortex.txt.gz:
HiC/GSE87112/Dorsolateral_Prefrontal_Cortex.txt.gz
HiC/GSE87112/Hippocampus.txt.gz
Roadmap – brain: E053:E054:E067:E068:E069:E070:E071:E072:E073:E074:E081:E082

##### Single cell RNA-sequencing data sets used in the cell-type specific analyses

PsychENCODE_Developmental
PsychENCODE_Adult
Allen_Human_LGN_level1
Allen_Human_LGN_level2
Allen_Human_MTG_level1
Allen_Human_MTG_level2
DroNc_Human_Hippocampus
GSE104276_Human_Prefrontal_cortex_all_ages
GSE104276_Human_Prefrontal_cortex_per_ages
GSE67835_Human_Cortex
GSE67835_Human_Cortex_woFetal
Linnarsson_GSE101601_Human_Temporal_cortex
Linnarsson_GSE76381_Human_Midbrain

### Supplementary Figures

##### Supplementary Figure 1. Heterogeneity across cohorts.

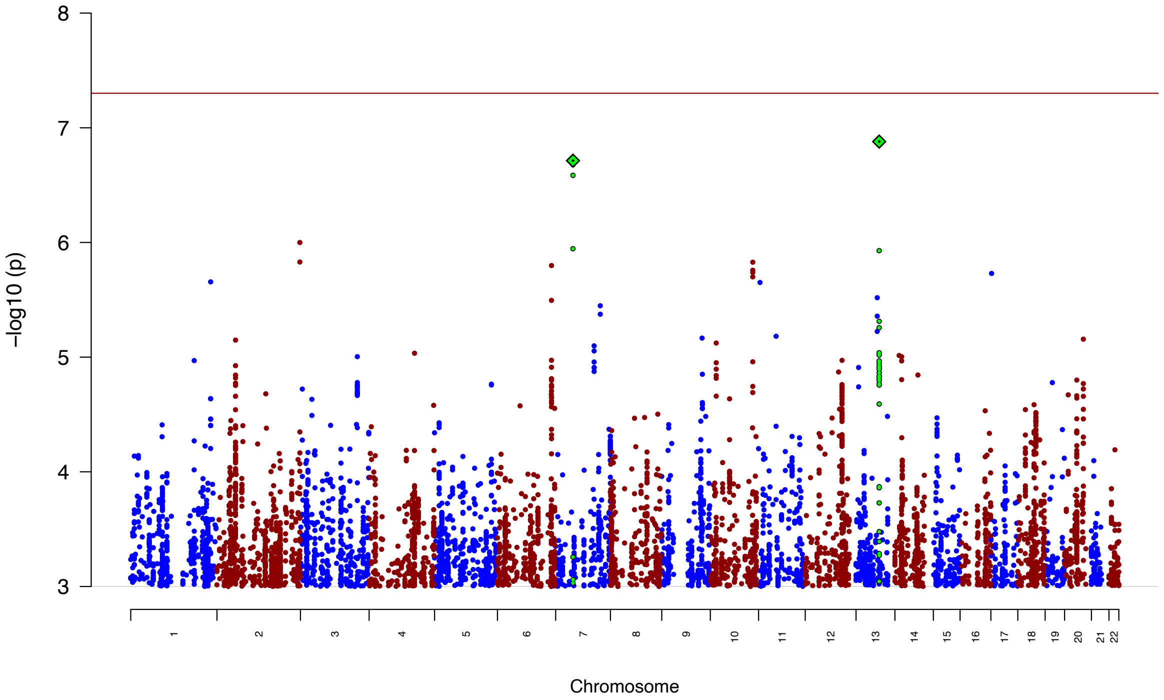


Test for heterogeneity across cohorts. The y-axis represents –log(*P*-values) from omnibus test of heterogeneity across cohorts (I squared statistic ($I^2$)) consisting of 38,691 cases and 186,843 controls. Red reference line indicates genome-wide significance threshold ($P = 5\times10^{-8}$).

#### Supplementary Figure 2. Quantile-quantile plot of GWAS meta-analysis of ADHD

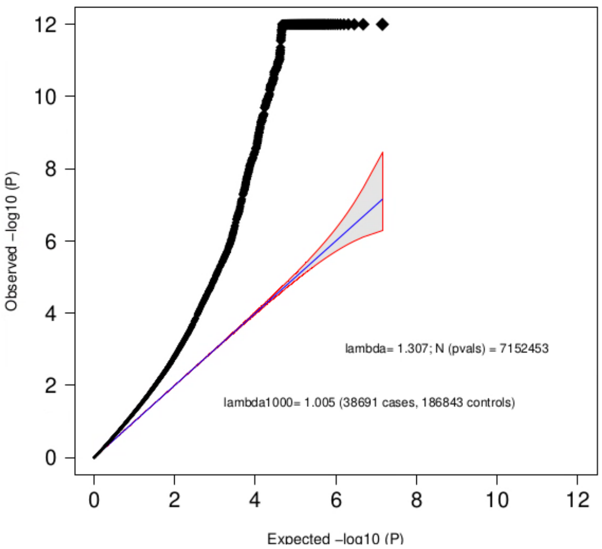


Quantile-quantile plot of the -log10 *P*-values from the GWAS meta-analyses of ADHD (38,691 cases and 186,843 controls) against expected -log(*P*-values). The blue line indicates the distribution under the null hypothesis and the corresponding standard error.

#### Supplementary Figure 3. Enrichment in ADHD risk genes among differentially Expressed Genes (DEG) in brain tissue

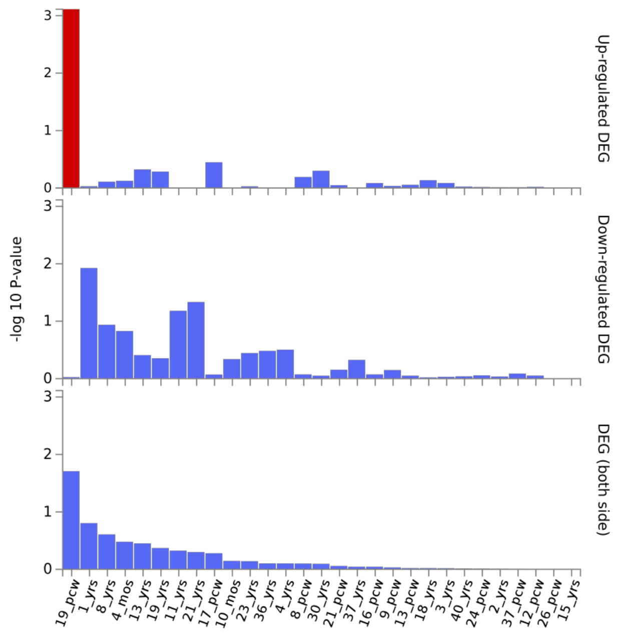


Results from hypergeometric test evaluating enrichment of the 76 ADHD risk genes among differentially Expressed Genes (DEG) in brain tissue representing different brain developmental stages in Brain Span. DEG sets were available in FUMA. Red bars indicate significant results.

#### Supplementary Figure 4. Enrichment analyses in previous GWAS studies

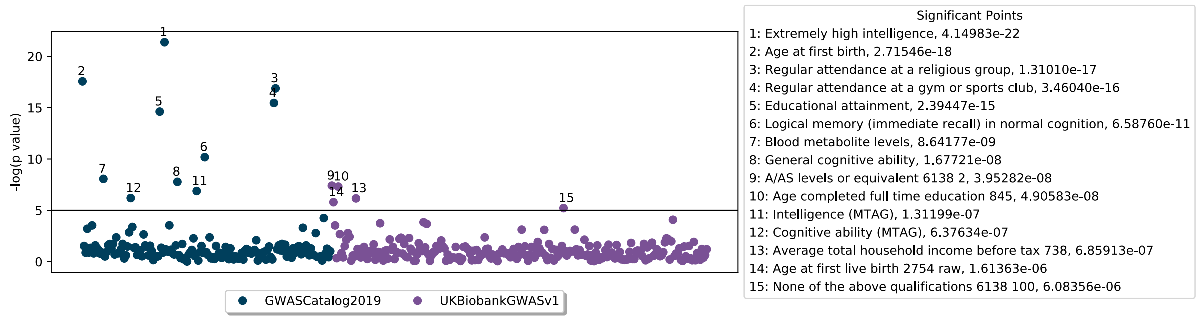


Results from enrichment analysis (hypergeometric test) of the 76 ADHD risk genes among sets of genes identified in the GWAS 2019 Catalog or UK Biobank. Significant phenotypes are listed at the right together with the enrichment *P*-value ($P < 0.00001$ are shown).

#### Supplementary Figure 5. Analysis of ADHD risk genes in SynGO

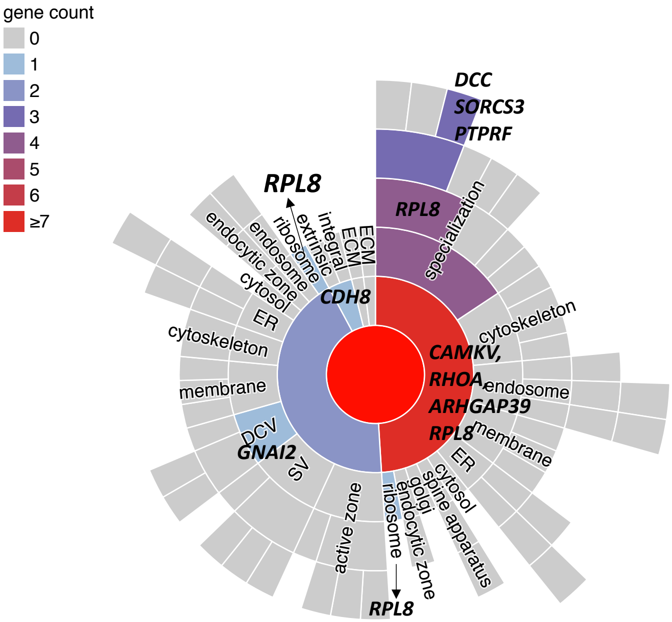


Results of SynGO ontology terms enrichment among the 76 ADHD risk genes as compared to the selected background brain expressed genes. Location of annotated genes with respect to cellular component i.e. where the gene product is active in the synapse. The gene count column shows the number of ADHD genes annotated in SynGO against a term or any of its child terms (more specialized terms). Unique genes for a term are specified. Gene counts (see color code in the bar at left) include unique genes and child terms. *ER* - endoplasmic reticulum; *SV* - synaptic vesicle; *DCV* - neuronal dense core vesicle; *ECM* - extra cellular matrix of synaptic cleft; *ECM* - perisynaptic extra cellular matrix.

#### Supplementary Figure 6. Miami plots of GWAS and TWAS results

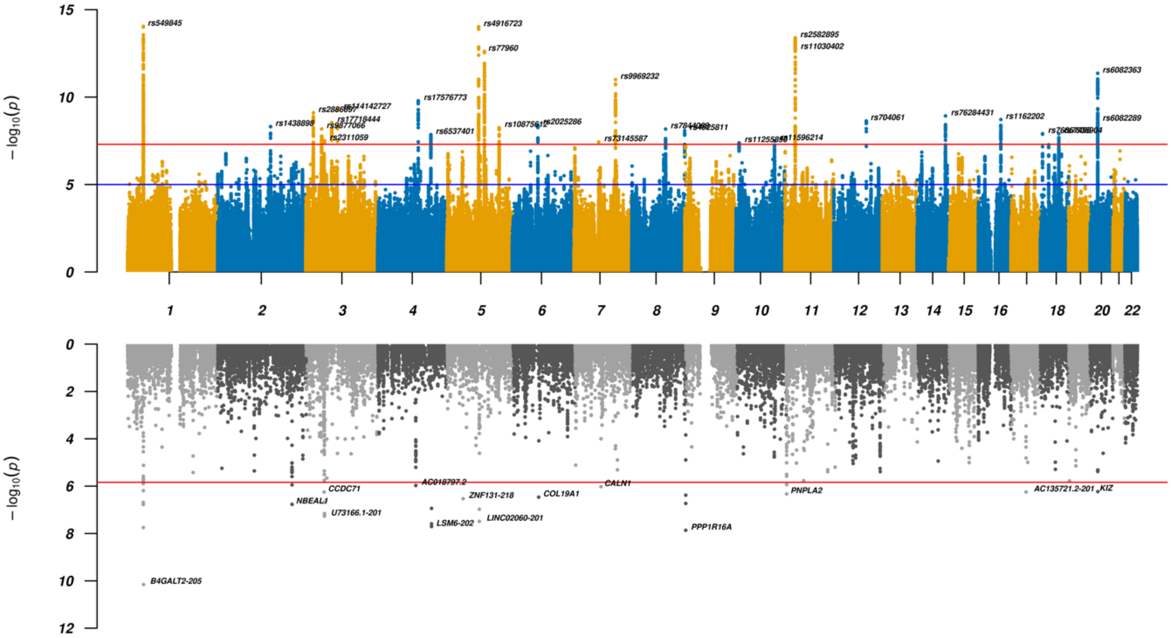


Results for GWAS (top panel) and TWAS results for DLPFC transcripts (bottom panel) for ADHD GWAS. In the top panel a blue line in the Manhattan plot indicates a P-value of $1\times10^{-5}$, a red line a P-value of $5\times10^{-8}$ (genome-wide significance). Each dot represents a tested SNP. In the bottom panel genes are represented by both gene expression and isoform expression (= features, represented by the dots). A red line indicates Bonferroni corrected genome-wide significance within analyses; $P < 1.44\times10^{-6}$; corresponding to Bonferroni correction of all the 34,646 features).

### Supplementary Figure 7.A-B. Regional Miami plots for ADHD GWAS and TWAS

#### A) Chromosome 1 locus - *B4GALT2-205* transcript

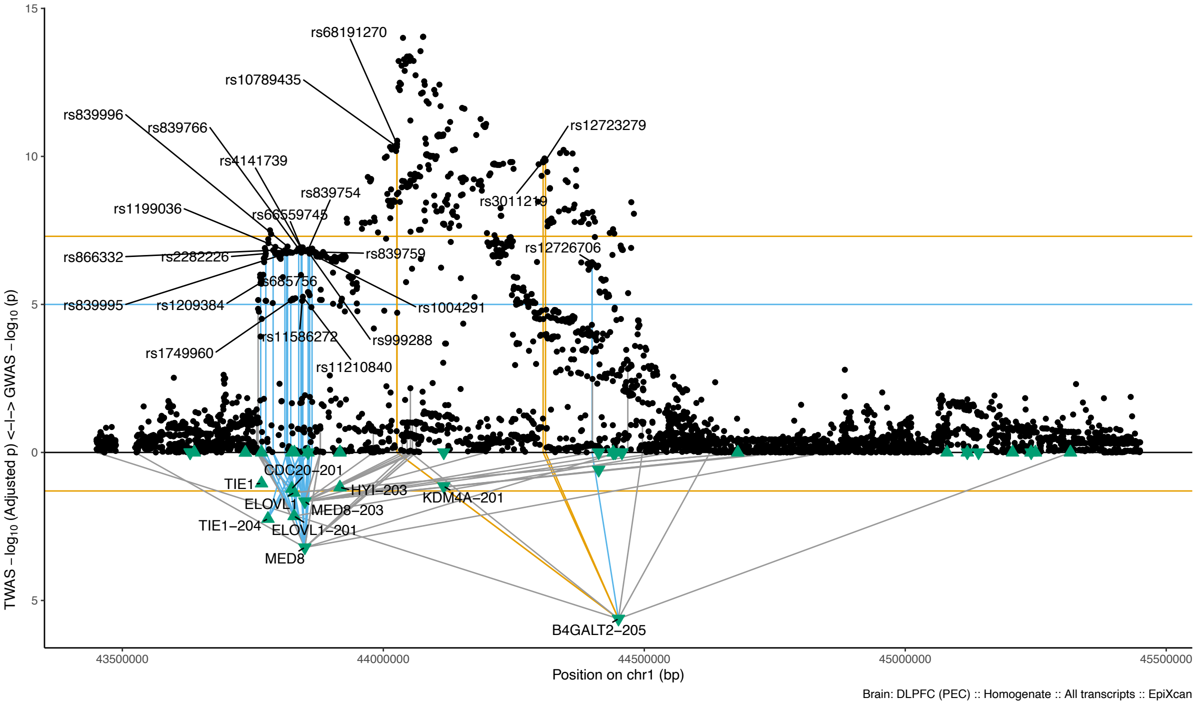


#### B) Chromosome 8 locus – *PPP1R16A*

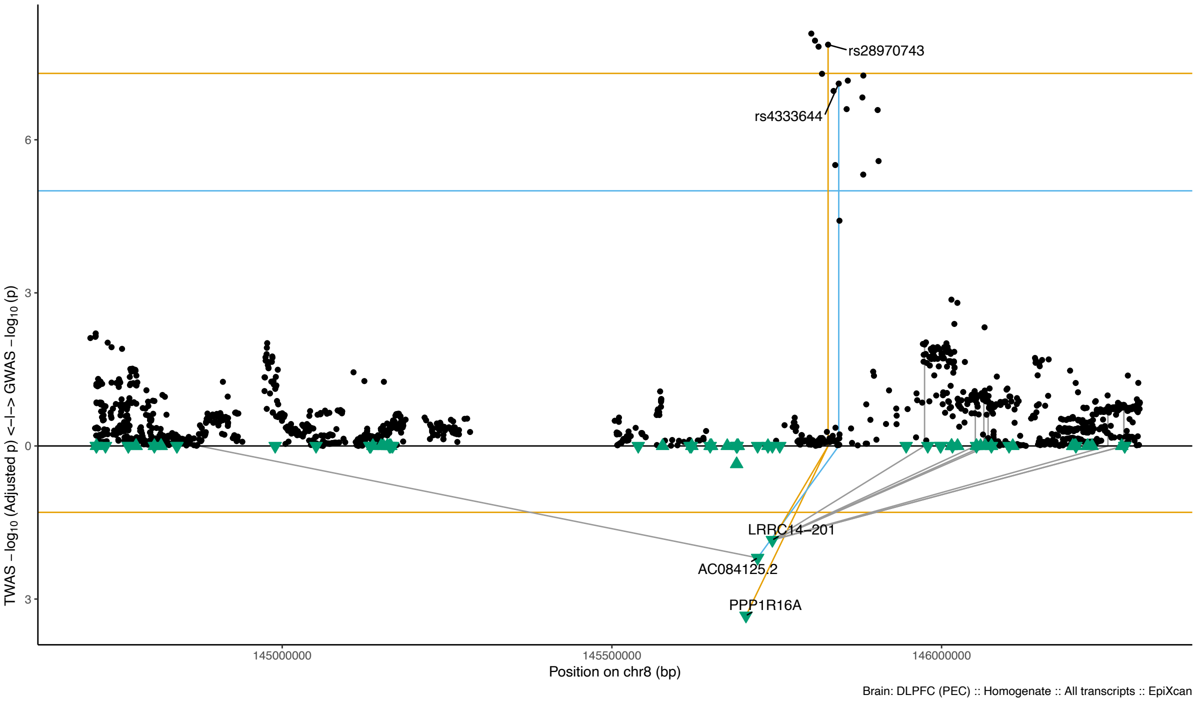

Regional Miami plots for ADHD GWAS meta-analysis corresponding to the genomic region of the respective transcript (1Mbp window from start site). Please also refer to Supplementary Table 8 and 9 for details. Top panel shows the GWAS results (black dots); blue line corresponds to $P = 1\times10^{-5}$, orange line to $P = 5\times10^{-8}$ (genome-wide significance). Bottom panel shows the TWAS results (green triangles facing upwards or downwards for a positive or negative z-score respectively; only the transcripts with Bonferroni-adjusted $P < 0.1$ are labelled for clarity) for different transcripts (genes are represented by both gene expression and isoform expression); orange line corresponds to Bonferroni-adjusted P = 0.05. Each transcript that is Bonferroni-significant in the region is connected with lines to the SNPs that contribute to its transcriptomic imputation model; lines are grey when the SNPs have a$P > 1\times10^{-5}$, blue when $P < 1\times10^{-5}$ but $> 5\times10^{-8}$ and orange when $P < 5\times10^{-8}$. The SNPs that are above the blue line and contribute to the transcriptomic imputation models of significant transcripts are labelled.

#### Supplementary Figure 8. Tissue specific gene expression of ADHD risk genes

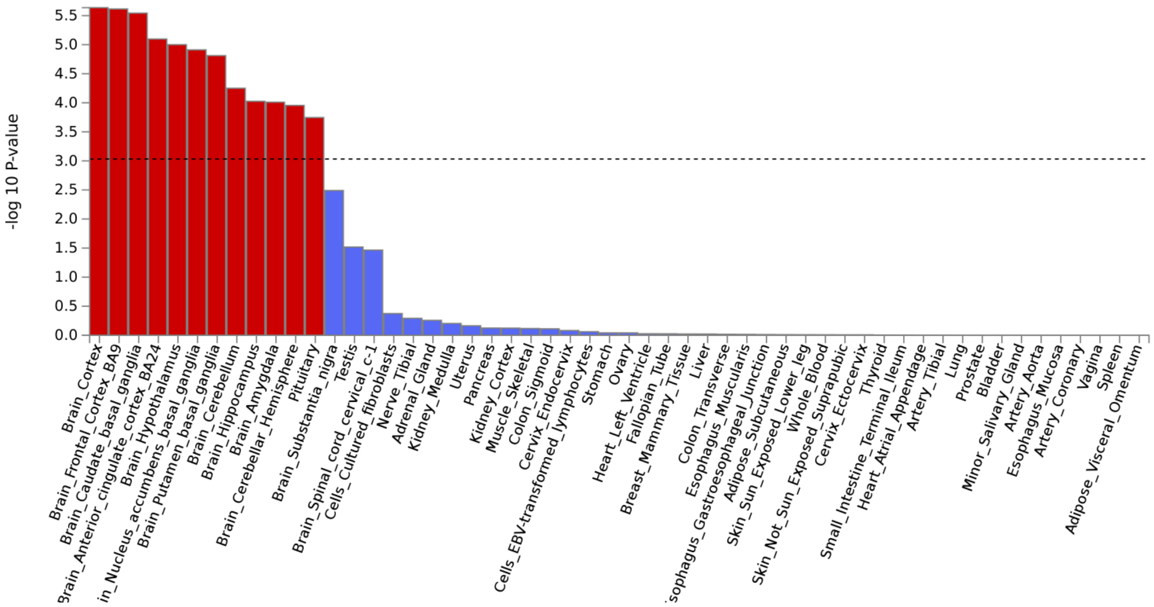


Results from MAGMA gene-property analysis of relationships between tissue specific gene expression profiles (GTEx v.7) and ADHD-gene associations. Red bars indicate significant results.

#### Supplementary Figure 9. Partitioned SNP-heritability across human cell types

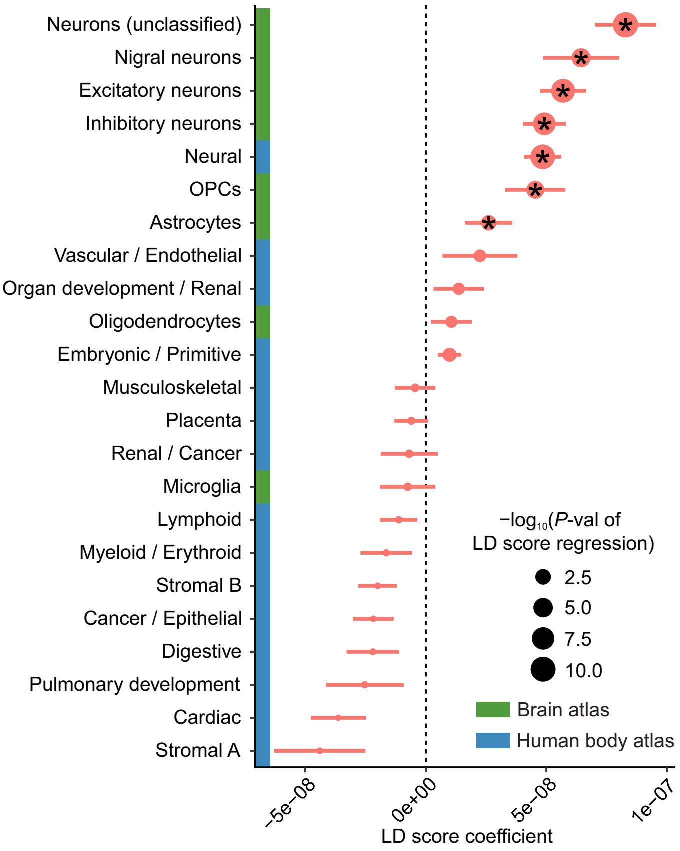


Enrichment of ADHD risk variants in genomic regions of open chromatin from DHS study (DNase I hypersensitive sites) in major human cell types (marked in blue on the y-axis) and from a scATAC-seq study (marked in green on the y-axis) single-cell assay for transposase accessible chromatin) of brain-specific cell types, using LD-score partitioned heritability approach. The positive LD score coefficient signifies enrichment in heritability (horizontal bares reflect standard error). Dot size reflects *P*-value of LD score regression, '*' denotes test-wide significant associations.

#### Supplementary Figure 10. Cell type specific expression of ADHD genes

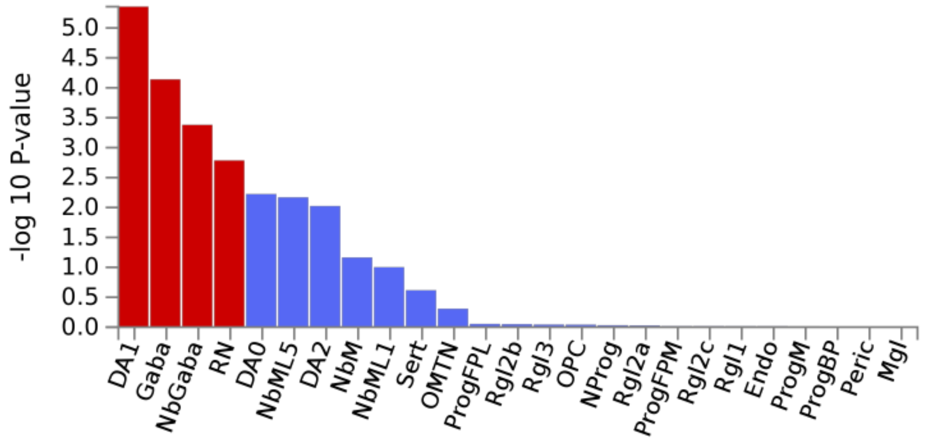


Results from magma-property analysis of "gene-ADHD association" and gene expression in Linnarsson midbrain single cell RNA-sequencing data. After correction for multiple testing across all datasets (red bars indicate significant results after this step) and subsequent systematic step-wise conditional analysis within the data set only dopaminergic DA1 neuron specific-expressed-genes were associated with ADHD (Supplementary Table 13). *DA0-2* - dopaminergic neurons; *Gaba* - GABAergic neurons; NbGaba - neuroblast gabaergic; *RN* - red nucleus*;* *Endo* - endothelial cells; *Mgl* - microglia; *NProg* - neuronal progenitor; *NbM* - medial neuroblast; *NbML1*- mediolateral neuroblasts; *OMTN* - oculomotor and trochlear nucleus; *OPC* - oligodendrocyte precursor cells. *Peric* - pericytes; *Prog* - progenitor medial floorplate (FPM), lateral floorplate (FPL), midline (M), basal plate (BP); *Rgl1-3* - radial glia-like cells; *Sert* – serotonergic.

#### Supplementary Figure 11. Cell type specific expression of ADHD genes

A) Chromosome 1 (*PPP1R16A*)

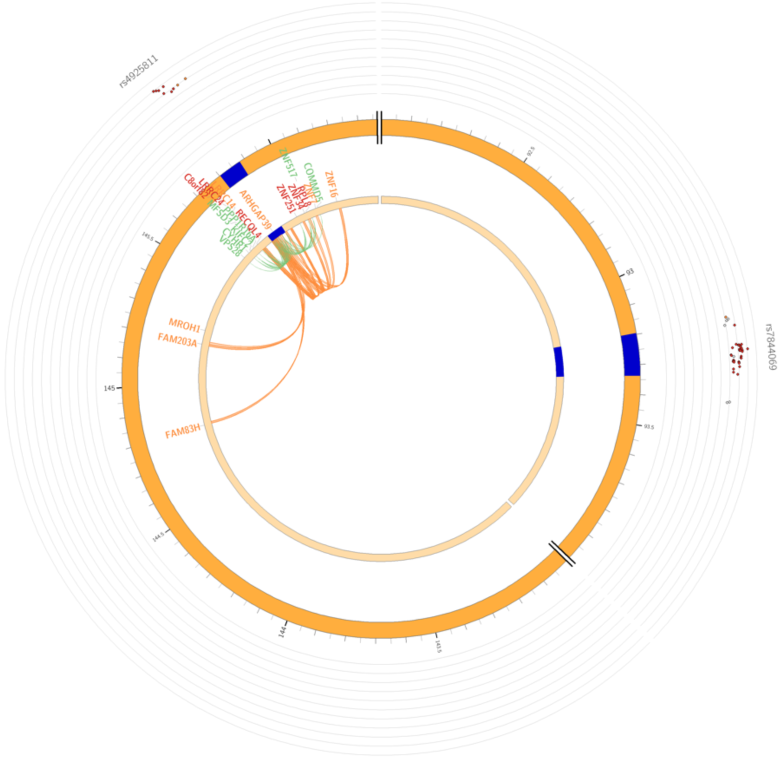


B) Chromosome 8 (*BAGALT2*)

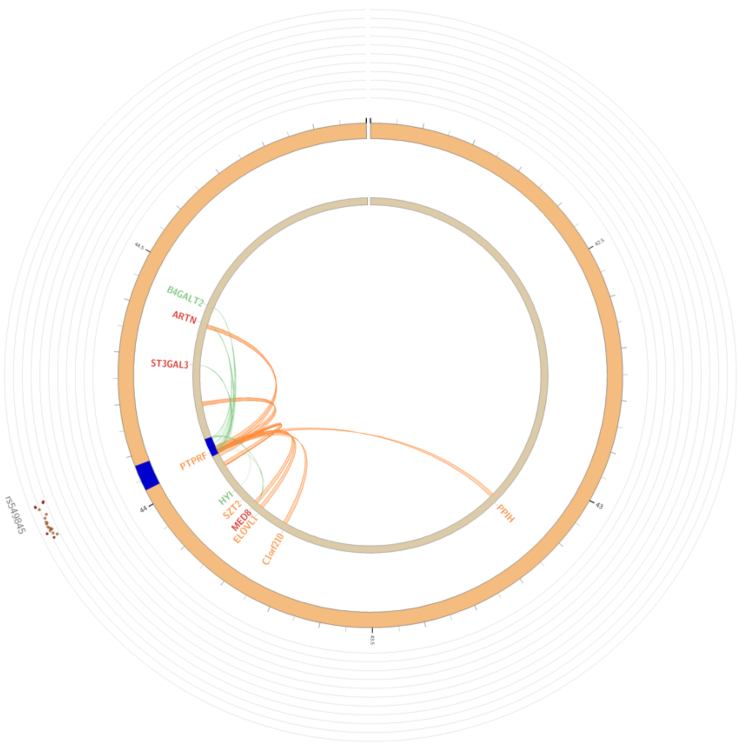

#### C) Chromosome 7 (*FOXP2*)

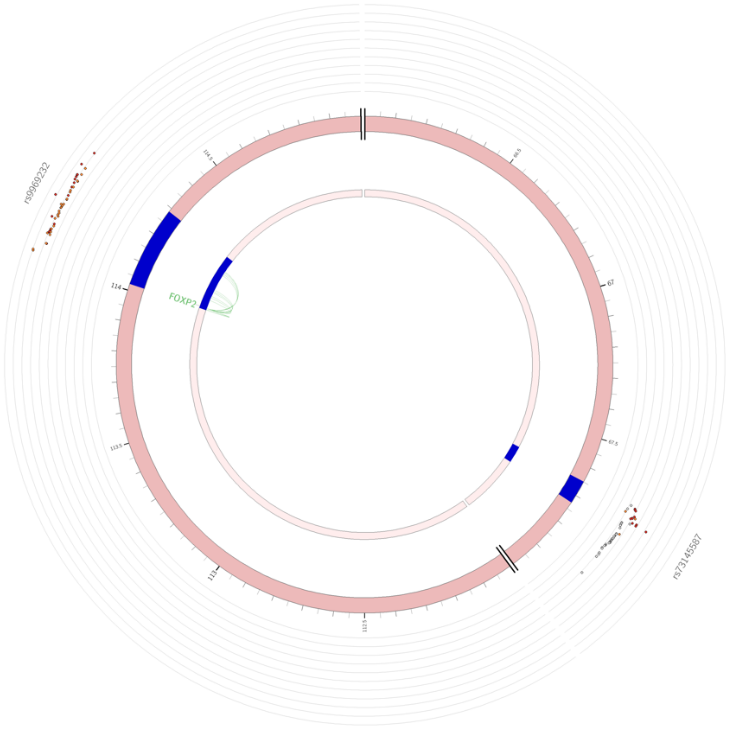


#### D) Chromosome 3 (*FOXP1*)

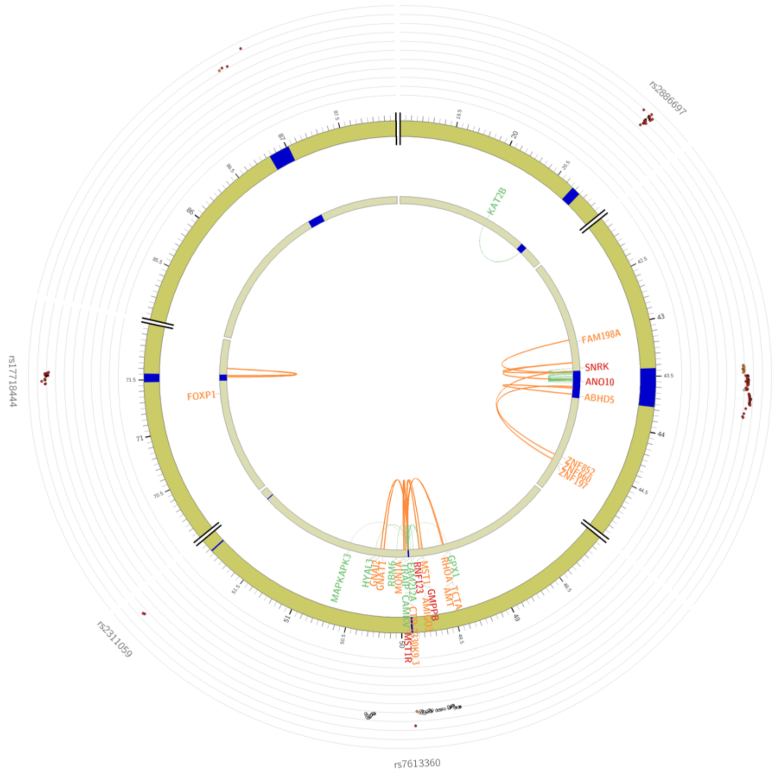

Circus plot: Association of credible variants in the ADHD GWAS meta-analysis in the most outer layer. SNPs in genomic risk loci are color-coded as a function of their maximum $r^2$ to one of the independent significant SNPs in the locus, as follows: red ($r^2 > 0.8$), orange ($r^2 > 0.6$), green ($r^2 > 0.4$) and blue ($r^2 > 0.2$). SNPs that are not in LD with any of the independent significant SNPs (with $r^2 \leq 0.2$) are grey. The rsID of the top SNPs in each risk locus are displayed in the most outer layer. Y-axis are ranged between 0 to the maximum -log10(*P*-value) of the SNPs.
Chromosome ring: The second layer. Genomic risk loci are highlighted in blue.
Mapped genes by chromatin interactions or eQTLs: Only mapped genes by either chromatin interaction and/or eQTLs are displayed. If the gene is mapped only by chromatin interactions or only by eQTLs, it is colored orange or green, respectively. When the gene is mapped by both, it is colored red.
Chromosome ring: The third layer. This is the same as second layer but without coordinates to make it easy to align position of genes with genomic coordinate.
Chromatin interaction links: Links colored orange are chromatin interactions. eQTL links: Links colored green are eQTLs.
